# Species-specific bacterial detector for fast pathogen diagnosis of severe pneumonia patients in the intensive care unit

**DOI:** 10.1101/2022.03.25.22272920

**Authors:** Yan Wang, Xiaohui Liang, Yuqian Jiang, Danjiang Dong, Cong Zhang, Tianqiang Song, Ming Chen, Yong You, Han Liu, Min Ge, Haibin Dai, Fengchan Xi, Wanqing Zhou, Jian-Qun Chen, Qiang Wang, Qihan Chen, Wenkui Yu

## Abstract

Rapid diagnosis of pathogens is the cornerstone of appropriate therapy and is also a great challenge to be overcome. Although NGS and some other PCR-based pathogen detection methods were applied to improve the speed and accuracy of clinical diagnosis, it was still a long way from the clinical needs of rapid and accurate diagnostic therapy in the intensive care unit (ICU). In this study, we aimed at developing a new rapid diagnostic tool, Species-Specific Bacterial Detector (SSBD), to evaluate the existence and quantification of 10 most usual pathogenic bacteria in ICU in 4 hours. Briefly, the species-specific genome fragments of each bacterium were identified by our algorithm using 1791 microbe genomes from 232 species and then used to combine with CRISPR/Cas12 to establish diagnosis tools. Based on the tests of 77 samples, SSBD demonstrated 100% sensitivity and 87% specificity compared with conventional culture test (CCT). Later on, an interventional random-grouped study was applied to evaluate the clinical benefits of SSBD. Briefly, SSBD demonstrated more accurate and faster diagnosis results and led to earlier antibiotics adjustment than CCT. Based on the results acquired by SSBD, cultivation results could deviate from the real pathogenic situation with polymicrobial infections. In addition, nosocomial infections were found widely in ICU, which should deserve more attention.

## 1. Introduction

Sepsis is associated with high morbidity and mortality [1]. Adequate antibiotic therapy in time could decrease mortality and reduce the length of stay in ICU for patients with sepsis or septic shock [2–5]. As reported in the previous study, the mortality rate of patients increased approximately 7.6% for every hour delayed [3]. Therefore, rapid diagnosis of pathogenic microorganisms is crucial for shortening the time of empirical antibiotic therapy and improving the prognosis of patients with sepsis.

Conventional culture test (CCT) is the most commonly used and golden standard identification method of pathogenic microorganisms in most countries. However, it showed two critical limitations: long time-consuming (2 to 5 days) and low sensitivity (30-50%), which limited the application of this method in the ICU [6, 7]. To overcome this bottleneck, several new tools were developed and showed significant improvement in time consumption and accuracy. Recently, next-generation sequencing (NGS) technology was applied to acquire the entire information of microorganisms and demonstrated great ability in diagnosing rare pathogens. However, the whole process still needs at least 2 days for the full diagnostic report with high cost [8, 9]. On the other hand, NGS provided too much information about microorganisms but only semi-quantification of pathogens, which was hard for most clinical doctors to extract the most important information to determine antibiotic usage. Other new emerging detection techniques designed by BioFire and Curetis are much superior in detection time than these above. However, its original principle was based on nucleotide diversity of conserved genes among species, which could not satisfy the application in the ICU due to potential false-positive results [10–12]. Therefore, a unique diagnosis tool aimed at faster and more accurate pathogen identification in the ICU was still a great challenge.

In this study, we aimed to design a simple and convenient diagnosis tool for sepsis patients in the ICU, which covered the most common pathogenic bacteria and completed the detection process in the shortest possible time with low cost and minimum instrument requirements. A clinical trial with two stages was applied to evaluate the accuracy of the tool and the clinical benefits.

## 2. Materials and Methods

### 2.1. Study design

The full study design was shown in **Fig 1**. In the discovery stage, we screened species specific DNA-tags of 10 epidemic pathogenic bacteria in the ICU. In the training stage, we optimized reaction conditions and sample preparation process, including detection concentration limitation, DNA purification, and incubation time of the CRISPR/Cas12a reaction. The finalized experiment operating procedure of SSBD was used in the subsequent stages (detailed protocol was shown in Supplementary Materials).

**Figure 1:**
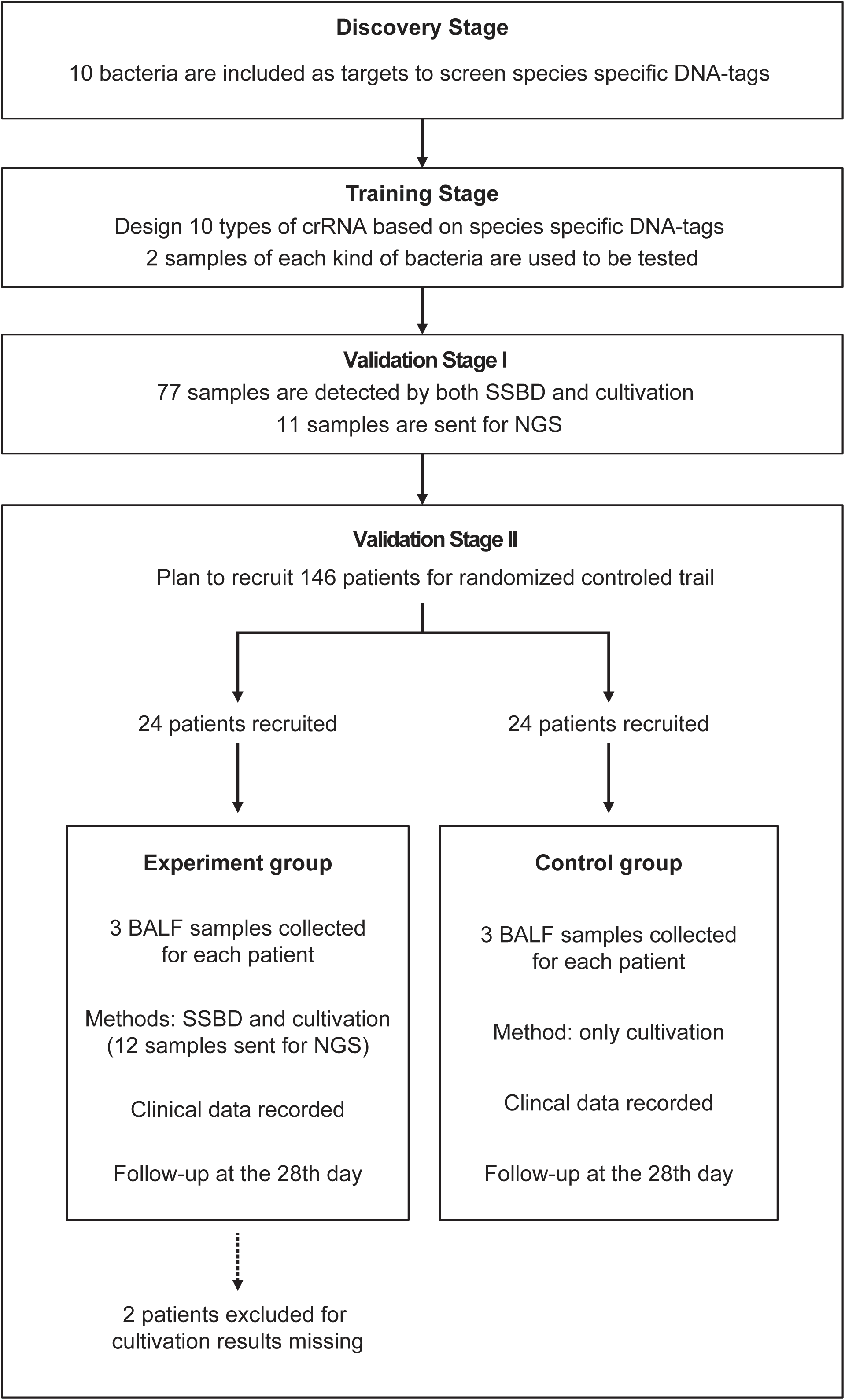
Study Design. This study contained four stages: discovery stage, training stage, validation stage I and validation stage II. All patients were from the Department of Critical Care Medicine, Nanjing Drum Tower Hospital. sPatients were randomly divided into two groups for the clinical trial.

In validation stage I, 77 specimens of bronchoalveolar lavage fluid (BALF) directly acquired from patients in ICU were finally detected by SSBD to confirm the specificity and sensitivity of SSBD compared to CCT results. Some of the samples were sent to NGS technology, providing additional references.

After the stability and accuracy of SSBD were thoroughly evaluated, the validation stage II, a preliminary clinical intervention experiment, was launched to verify the clinical application of the SSBD.

### 2.2. Ethics Approval

We acquired the ethics approval (2019-197-01) from the ethics committee of Nanjing Drum Tower Hospital Affiliated to Nanjing University Medical School in July 2019, registered and posted the complete research protocol, informed consent, subject materials, case report form, researcher manual, the introduction of main researchers and other information in Chinese. Later on, this study was registered in English at ClinicalTrilas.gov (NCT04178382) in November 2019.

### 2.3. Screening species-specific DNA tags

We designed a process to find the species-specific DNA tags according to the basic principle, intraspecies-conserved and interspecies-specific sequences (illustrated in **Fig 2A**). 1791 high-quality genomes of 232 microorganism species from the public databases were included in the screening process. To accelerate the screening process, we developed a linear comparison algorithm instead of comparing every two genomes, which could save more than 90% of calculation time cost (**S1 Fig**). 10 species of bacteria were selected as targets for subsequent detecting process, including *Acinetobacter baumannii* (*A. baumannii*), *Escherichia coli* (*E. coli*), *Klebsiella pneumoniae* (*K. pneumoniae*), *Pseudomonas aeruginosa* (*P. aeruginosa*), *Stenotrophomonas maltophilia* (*S. maltophilia*), *Staphylococcus aureus* (*S. aureus*), *Staphylococcus epidermidis* (*S. epidermidis*), *Staphylococcus capitis* (*S. capitis*), *Enterococcus faecalis* (*E. faecalis*) and *Enterococcus faecium* (*E. faecium*). Then we designed different DNA primers targeting selected species-specific DNA tags from each species (**S1 and S2 Table**).

**Figure 2:**
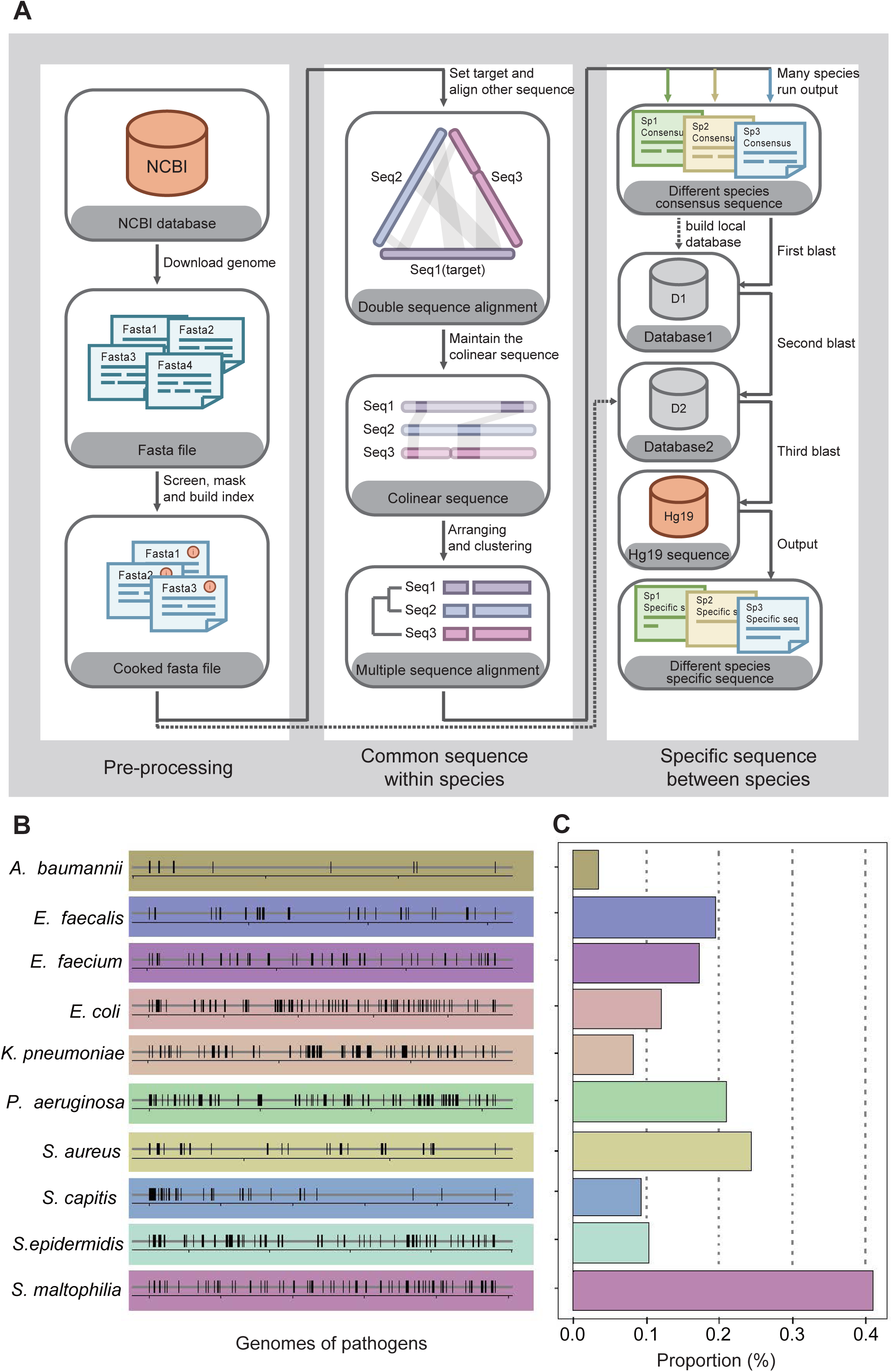
Screening workflow and statistics of species-specific DNA-tags. A. Schematic diagram of screening species-specific DNA-tags. B. Genomic distribution of species-specific DNA-tags in 10 bacteria. C. Genomic proportion of species-specific DNA-tags in 10 bacteria.

To evaluate our primers’ specificity in identifying species, we chose *S. aureus* and *S. epidermidis* from the same genus as our cross-validated target species. We extracted DNA sequences of the *S. aureus* and the *S. epidermidis* amplified by primers used in FilmArray Pneumonia Panel developed by BioFire and in our protocol, which were acquired from NCBI Reference Prokaryotic Representative genomes. We then aligned *S. aureus*-specific DNA sequences with the representative genome of *S. epidermidis* using blast to search the most similar DNA sequences. In the FilmArray Pneumonia Panel, DNA amplified sequences from the *S. aureus* and the *S. epidermidis* were aligned to each other (two different gene regions, *rpoB* and *gyrB*, were used to separate two species).

### 2.4. Patients

Patients admitted to ICUs and diagnosed with severe pneumonia were recruited from Aug 27, 2019. The recruit criteria for patients were: (1) age ≥ 18 years; (2) had artificial airway and expected to retain for more than 48 hours; (3) clinically diagnosed as pneumonia, and the microbiology of etiology was unclear; (4) signed informed consent;

(5) the expected length of staying in ICU was more than 3 days. According to previous mortality acquired from the adequate anti-infective group, the sample size calculation (two-group rate) for patients was done, and a sample size of 73 patients in each group was needed.

### 2.5. Clinical outcomes

BALFs were obtained from all the patients from 2 groups on day 1, day 3-5, and day 7-10 after recruitment and were sent directly to the hospital diagnostic microbiology laboratory for CCT and susceptibility testing. BALFs from patients of the experiment group were also sent for SSBD tests immediately after sampling. Other clinical records included blood routine tests, CRP and PCT examinations.

All enrolled patients received primary empirical antibiotic therapy. Once the SSBD results of the patients in the experiment group were obtained, the decisions about whether antibiotics were adjusted or not were made by two senior doctors according to the SSBD results and other clinical information. While in the control group, adjustment depended on conventional culture results and clinical data. Patient demographics and other vital clinical parameters were recorded. Acute Physiology and Chronic Health Evaluation II (APACHE II) scores and Sequential Organ Failure Assessment (SOFA) scores were calculated and recorded for patients on days 1, 3, 7, 10 and 14 to assess their disease severity and organ function.

### 2.6. Statistical analysis

The number of improved patients on different clinical indicators of different days was calculated and tested by Fisher’s exact test. APACHE II scores and SOFA scores were tested as a series by two-way ANOVA. Different clinical outcomes and TTAT were tested by the Mann-Whitney test.

### 2.6. Funding support

This study was funded by National Natural Science Foundation of China. The National Key Scientific Instrument and Equipment Development Project. Project number: 81927808.

## 3. Results

### 3.1. The identification of species-specific DNA fragments

The first step to identify pathogenic bacteria was to figure out the specific genome information of each species. Unlike viruses or animals, bacteria were quite similar between close-related species but sometimes quite different among different strains of one species due to fast evolution and horizontal gene transfer [13–15]. Therefore, the widely-used method to identify bacteria with conserved genes may not be a good choice [16, 17]. We developed an innovative algorithm and designed a workflow to figure out the best DNA tag for each species for diagnostic application based on 1791 microbe genomes from 232 species (**Fig 2A**). The details could be found in the Supplementary Materials.

We started from 10 common bacteria contributing to sepsis infection as the initial panel according to local epidemic data from ICU of Drum Tower hospital and previous studies about pathogens in ICU (**S2 Fig**) [18, 19]. To our surprise, bacteria-specific DNA sequences showed a random distribution and turned out to be only 0.3%-4.1% in the whole genomes of 10 bacteria (**Fig 2B and 2C**). Considering the application scenario of ICU with only basic instruments, PCR+CRISPR/Cas12a system was chosen for the following detection. Based on the identified species-specific DNA fragments, related primers and crRNAs (CRISPR RNA) were designed according to each species (**S1 and S2 Table**).

### 3.2. The establishment of species-specific bacteria detection tool

Briefly, CRISPR/Cas12a with designed crRNA could be activated by its target, which could be told by whether the reporter probe was cleaved and demonstrated signal as previously reported [20].

To optimize the working conditions of the detection tool in ICU, multiple experiments were applied to optimize the sample preparation and detection process. With the gradient concentration of DNA templates, we confirmed that the lowest detection limit was 10^−15^ M with PCR amplification and 10^−8^M without amplification step (**Fig 3B**), which was consistent with previous studies [21]. In addition, 30 minutes’ incubation of CRISPR/Cas12a with PCR products was enough to demonstrate signals (**Fig 3B**). An additional purification step right after PCR amplification appeared unnecessary to acquire the positive result but helpful for weaker signal (**S3A Fig**). In addition, the comparison of CRISPR/Cas incubation duration confirmed that fluorescence value showed a significant difference from 5 minutes and reached its maximum after 30 minutes compared to the negative control (**S3B Fig**).

**Figure 3:**
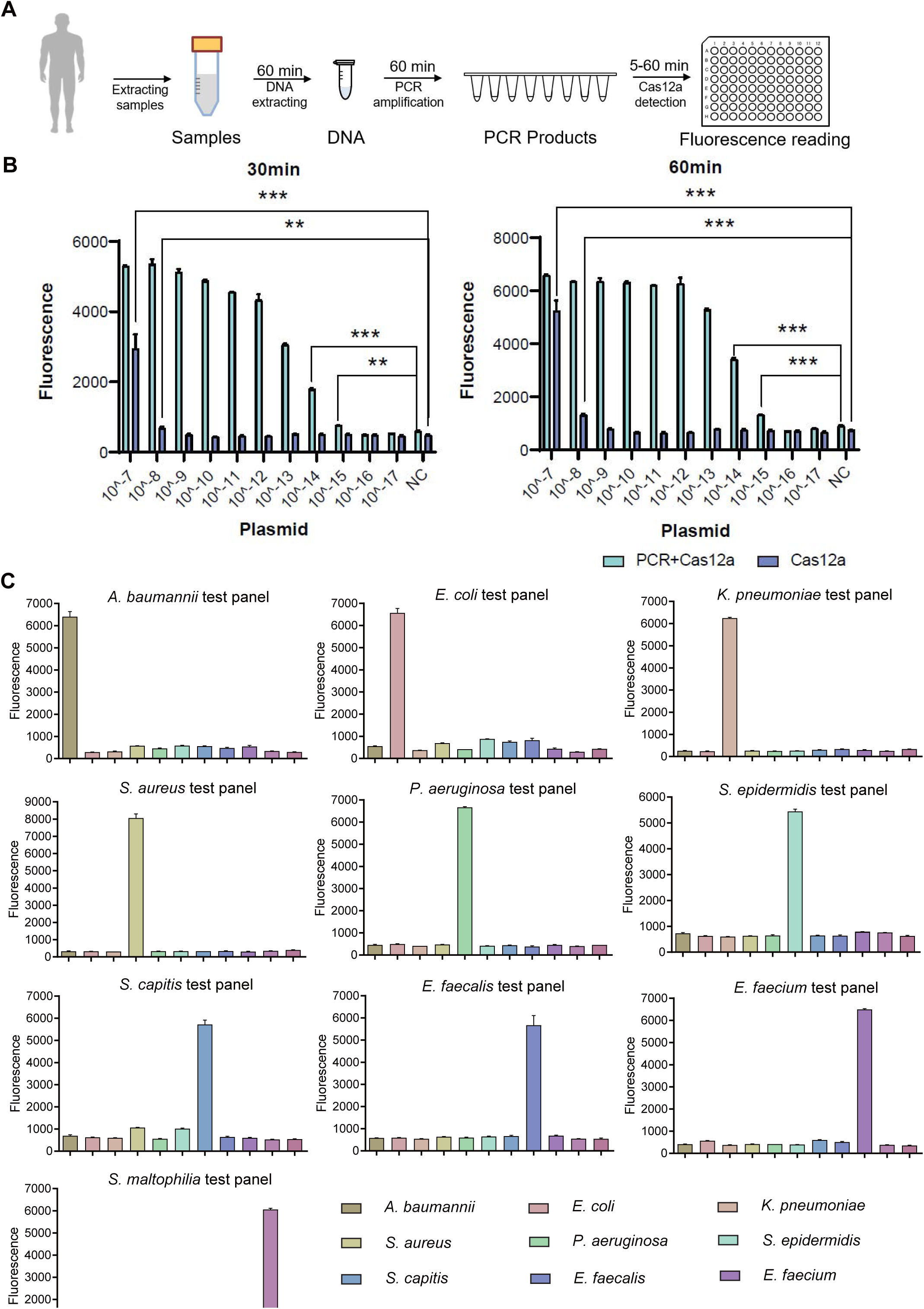
SSBD development and effectiveness validation. A. SSBD workflow for clinical validation stages. B. Cas12a and Cas12a-after-PCR detection of different concentrations and reaction times including 30 minutes (left) and 60 minutes (right). Blue bars indicated the Cas12a-after-PCR test. Brown bars indicated Cas12a test only. The concentration gradient of pGL3 plasmid from 10^−17^ M-10^−7^M was established as the test group. NC stood for the fluorescence values of PCR products of using DEPC-H2O as input. Each group had three repeats. Error bars indicated mean±SEM of fluorescence value. ** indicated p value<0.01 and *** indicated p value<0.001 of unpaired t-test. C. SSBD results of 10 pathogenic bacteria. Every test panel for each of 10 bacteria was used to detect genome DNA samples of 10 bacteria by SSBD. NC stood for the fluorescence values of PCR products of using DEPC-H2O as input. Each group had three repeats. Error bars indicated mean±SEM of fluorescence value.

To confirm the primary behavior of SSBD, two clinical strains separated from different patients for each of 10 selected bacteria species were collected and tested by SSBD as the positive control, which showed clear positive results (**S3C Fig**). To further confirm the specificity of SSBD, each bacteria strain was tested by 10 SSBD test panels targeting different bacteria. Compared to negative control, only SSBD targeting the tested bacteria showed a positive result, which confirmed its high specificity (**Fig 3C**). Putting these results together, a standard operating procedure was finally established for the following validation stages (**Fig 3A**), which was capable of providing the information about the ten most common pathogenic bacteria in ICU. Since this method was a quite fast and <u>s</u>pecies-<u>s</u>pecific <u>b</u>acteria <u>d</u>etection tool, we named it SSBD.

### 3.3. The accuracy and clinical benefits of SSBD

We started our study with validation stage I, which was a non-intervention study with 77 samples of BALF extracted from patients. Samples were detected both by SSBD and CCT, and the results were compared (raw detection results were shown in **S3 Table**). Generally, 5 of 10 selected bacteria were detected by both tests, including *A. baumannii, K. pneumoniae, P. aeruginosa, S. aureus* and *S. maltophilia*. SSBD could detect those 5 bacteria separately with 100% sensitivity and over 87% specificity, which were calculated by the results of CCT as golden standard (**Fig 4A**). The other 5 bacteria were detected by SSBD but not CCT, including *E. coli, S. epidermidis, S. capitis, E. faecalis* and *E. faecium*. To further evaluate the results, 11 samples among all 77 samples were collected randomly and sent for NGS detection, which confirmed that SSBD and NGS demonstrated quite high similarities in results (**Fig 4A**).

**Figure 4:**
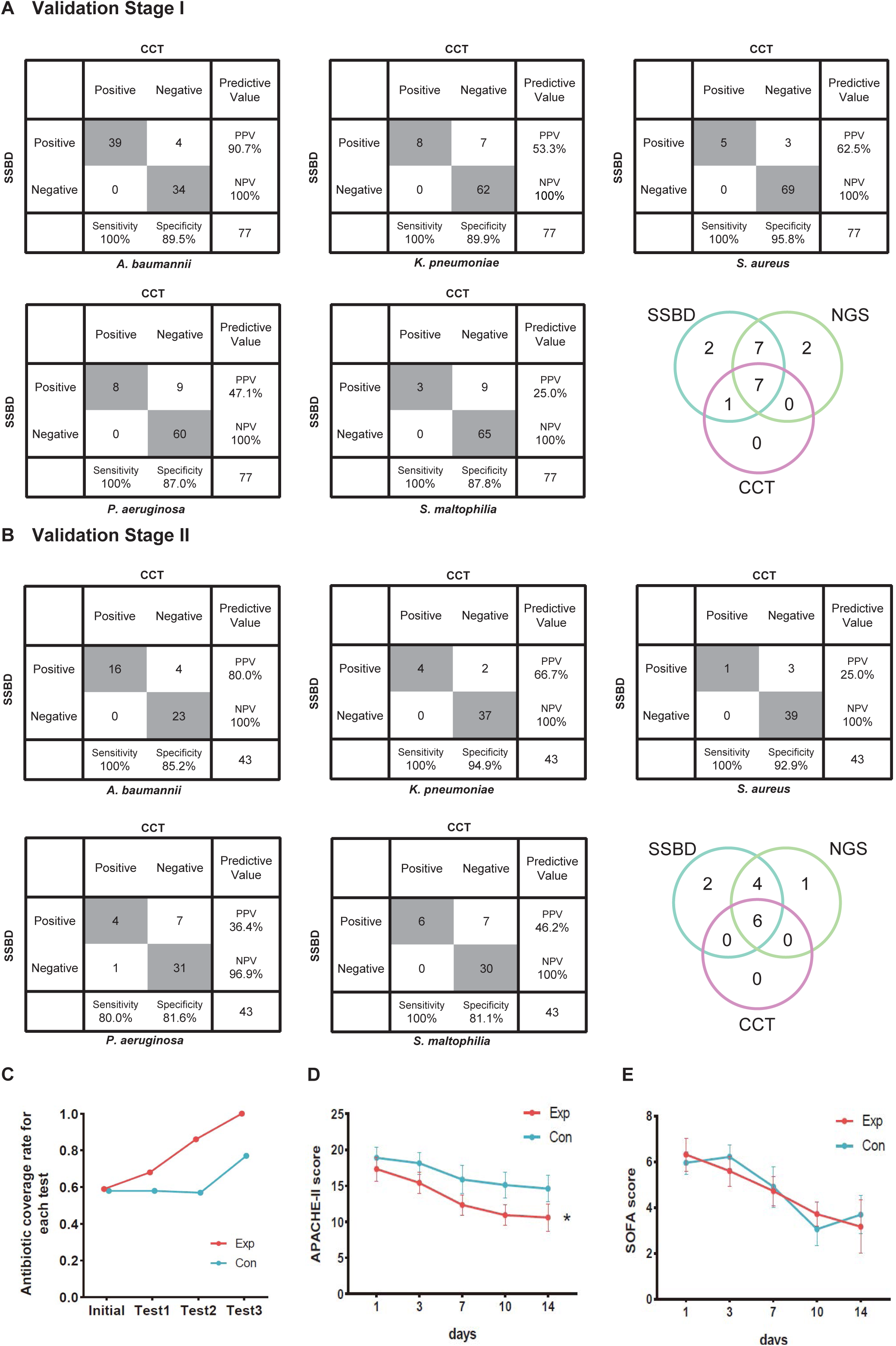
Statistical analysis of test results and clinical outcomes in the two validation stages. A. Cross-tables for 5 of 10 bacteria by both SSBD and CCT and comparative analysis of SSBD, CCT and NGS results in the validation stage I. B. Cross-tables for 5 of 10 bacteria by both SSBD and CCT and comparative analysis of SSBD, CCT and NGS results in the validation stage II. C. Antibiotics coverage rate of each test in the two groups. Exp meant the experimental group, and Con meant the control group. Test 1: Day 1. Test 2: Day 3-5. Test 3: Day 7+. Raw antibiotics coverage results of each patient were available in S2B Fig. Detailed judging guidelines were shown in Supplementary Material. D and E. Line charts for APACHE II and SOFA scores, respectively. Error bars indicated mean±SEM of scores of all the recorded patients. * indicated a significant difference between the two groups using two-way ANOVA.

Based on these accurate results, we started the validation stage II, which was an intervention study aiming to evaluate the clinical benefits of SSBD compared to the current diagnosis and treatment strategy in ICU. Although the study was paused due to the outbreak of SARS-CoV2, 22 patients were recruited into the experiment group and 24 patients into the control group. The baseline characteristics had no significant difference except ages (**S5 Table**).

We finally got 57 BALF results tested by SSBD, which included 43 results that also had CCT results among them in the experiment group. While in the control group, we got 63 samples tested only by CCT. In the experiment group, 47 samples were positive among 57 samples tested by SSBD, while 28 samples were positive among 43 samples tested by CCT (raw detection results were shown in **S4 Table**). In the control group, 41 samples showed positive among 63 samples. It was shown that SSBD could detect each bacterium with similar high sensitivity and specificity in validation stage II (**Fig 4B**). Consistent with the local epidemic data, the most frequent occurrence was *A. baumannii* (**Fig 4B**). 12 of the 57 samples were also sent for the NGS technology, and the SSBD also showed high consistency between those two methods compared to CCT (**Fig 4B**).

To explore clinical benefits with the help of SSBD, effective antibiotic coverage rate, APACHE II scores and SOFA scores were calculated and compared to evaluate the rationalization of antibiotic therapy and patients’ disease severity and organ function status in the two groups (**Fig 4C-4E**). Effective antibiotic coverage rates for each test were significantly higher in the experimental group than those in the control group in three tests (**Fig 4C**). The definition of antibiotic coverage and the original calculation results were shown in **S4A-B Fig**. APACHE II scores were significantly lower in the experimental group than those in the control group after day 1 (p=0.0035, two-way ANOVA); the separation between two groups of patients increased progressively until day 14 (**Fig 4D**). SOFA scores showed no difference between the groups (p=0.8918, two-way ANOVA) (**Fig 4E**). Other clinical outcomes showed no significant difference in both groups, such as time of ventilation, shock, 28-day mortality and the numbers of antibiotic-associated diarrhea (**S6 Table**).

### 3.4. Polymicrobial infection and nosocomial events observed by SSBD

Based on the previous studies, CCT had defects in the evaluation of polymicrobial infection events due to the limitations of its technology [22]. Therefore, we tried to evaluate whether SSBD demonstrated better performance with polymicrobial infection. Here, we defined situations of infection with more than one pathogenic microorganism as polymicrobial infection events to assess the performance based on the results of both methods. From the results, the detection rate of polymicrobial infection events by SSBD was 41.8% (55/134) in two validation stages, which was significantly higher than 11.7% (14/120) of CCT (**Fig 5A**). To confirmed the reliability of the extra detected polymicrobial infection events by SSBD, 6 samples were sent for NGS, which demonstrated the same results as SSBD (**Fig 5B**).

**Figure 5:**
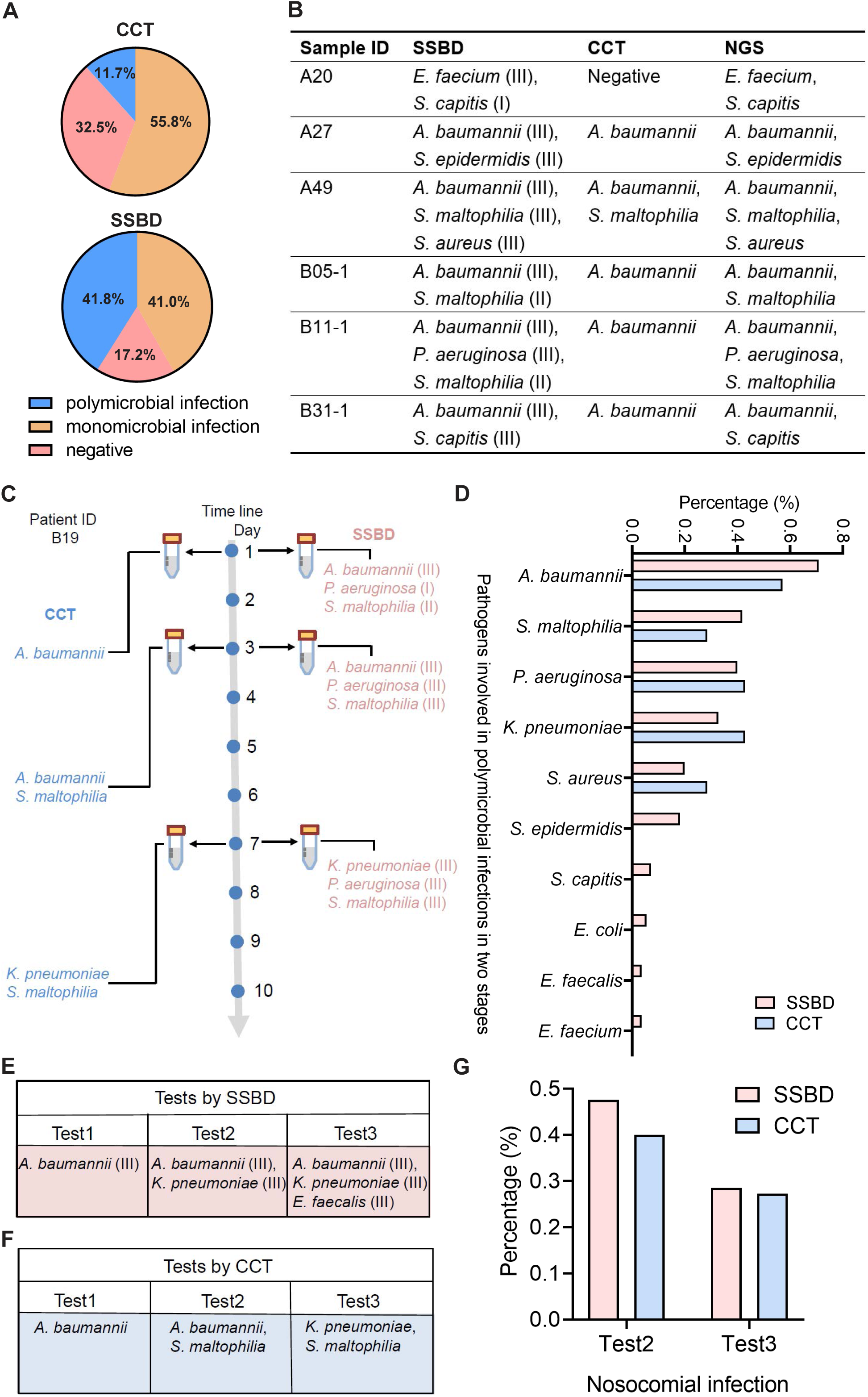
Statistical analysis of polymicrobial infection and nosocomial infection in the two validation stages. A. Statistics of pathogenic infection status of BALF samples in the two validation stages. B. Verification from NGS results for 6 samples identified as polymicrobial infection by SSBD but not CCT. C. Case study of polymicrobial infection detected by SSBD and CCT. E. Statistics of pathogens involved in polymicrobial infections in the two stages. F. Case study of nosocomial infection identified by SSBD. G. Case study of nosocomial infection identified by CCT. H. Percentage of nosocomial infection identified by SSBD and CCT.

Since both SSBD and NGS were based on target DNA, we wanted to confirm if some polymicrobial infection events were “false positive” and caused by dead bacteria. Here, we showed patient B19 as an example, who received three times tests at day 1, 3 and 7 by both CCT and SSBD. Based on the results, *S. maltophilia* was detected as level II in test1 with SSBD but not CCT. Later on, *S. maltophilia* was detected by CCT in test2 as well with few days’ development from level II to level III based on result of SSBD, which means SSBD discovered the true polymicrobial infection event earlier than CCT (**Fig 5C**). From the aspect of pathogen species participated in polymicrobial infection events, both methods demonstrated similar results with *A. baumannii, S. maltophilia, P. aeruginosa, K. pneumoniae* and *S. aureus* in top 5 (**Fig 5D**), which were consistent with the frequency of pathogens in ICU [18].

Hospital infections, also known as nosocomial infections, are an important factor in the incidence rate and mortality of ICU patients with severe pneumonia [23]. Since CCT has a long delay in clinical feedback of pathogenic results, there is no effective monitoring method in clinical practice. Here, we tried to evaluate nosocomial infections based on the test results. We defined a case as a nosocomial infection event if a pathogenic bacterium was newly detected in the current time point but not before. For example, B17 (*K. pneumoniae* at test 2, *E. faecalis* at test 3) and B19 (*S. maltophilia* at test 2, *K. pneumoniae* at test 3) patients were discovered as nosocomial infection cases for SSBD and CCT (**Fig 5E and 5F**). Based on the results of SSBD, 47.6% (10/21) of patients had nosocomial infections at the test 2, and 28.6% (4/14) of patients had nosocomial infections at the test 3. Similarly, 40% (4/10) of patients were identified as nosocomial infections by CCT at test 2, and 27.3% (3/11) of patients were identified as nosocomial infections at test 3 (**Fig 5G**).

## 4. Discussion

In this study, we developed a rapid bacteria detection technique based on CRISPR/Cas12a using species-specific DNA tags and detected common bacteria taken from pneumonia in 4 hours with 100% sensitivity and over 87% specificity in the validation stage I. Currently, there are already some market-oriented detection technologies for pneumonia patients, such as FilmArray Pneumonia Panel by BioFire and Curetis Unyvero system, which also could detect microorganisms in several hours [12]. Sequences used by FilmArray Pneumonia Panel from two gene regions had highly similar DNA sequences in the *S. epidermidis* representative genome (E-value=5e-40, *rpoB*; E-value=8e-39, *gyrB*), which could interfere with pathogen identification between species from the same genus. It was ideal for early and rapid screening of infectious diseases but was not applicable in the ICU, considering the complexity and urgency of infection events within the ICU. We have adopted a completely different strategy from the existing methods, getting specific gene regions from species for further test using our developed bioinformatics workflow and algorithm. It was shown that our sequences used for *S. aureus* diagnosis had no similar fragments in *S. epidermidis*, which avoided distinguishing different species by gene diversity. It was likely to get the species-specific DNA tags from such amount genomes when aligned bacterial genomes with each other but consuming computational cost. We optimized calculation processes by rescheduling steps and then made it possible for us to acquire species-specific DNA regions after shortening time to a range bearable.

NGS technology is useful in species identification and also shows its advantages in clinical diagnosis. It is valuable to detect uncommon pathogens because of its unique capability in detecting multiple agents across the full microbial spectrum contributing to disease and has already been developed as a new detection platform [9]. However, in the majority of cases of common pathogens, redundant microorganism results were probably unhelpful to the anti-infection regimen. In addition, the high cost and relatively long turnaround time prevent its widespread application, especially in the ICU circumstance. Therefore, our SSBD method seemed more advantageous in time-consuming and information effectiveness than other mentioned methods, especially when we could quantify bacterial load based on fluorescence intensity for better antibiotic therapy strategy. CRISPR/Cas12a and qPCR are both quantitative methods, but CRISPR/Cas12a shows its robustness and lower equipment requirement, which satisfied our needs for most of the ICU. There are still several challenges in implementing POCT in developing countries, especially the qPCR/POCT system, which will be an alternative.

The results of SSBD demonstrated high sensitivity and specificity. However, we discovered several “false positive” results compared to CCT, which might be caused by two reasons: 1) The low bacterial load of the patient sample was probably not enough or needed much longer time than expected to be cultivated. SSBD provided a lower threshold of detection (10^−15^ M) than CCT, which could detect pathogens that even existed in trace amounts which unable to be cultivated. In our study, the fluorescence intensity obtained from SSBD was divided into three intervals (level I: 10^−15^-10^−14^ M, level II: 10^−14^ M-10^−13^ M, level III: over 10^−13^ M), representing the different strengths of bacteria (roughly equivalent to bacteria amounts according to our lowest detection thresholds, dividing details in Supplementary Method SSBD diagnostic report and **S4A Fig**). All false-positive results were calculated on the count of species and strengths, mostly belonging to the level I or II (**S5 Fig**). Considering most of those false positive samples were also validated by NGS technology, it suggested that some pathogens might be missed in the CCT results. 2) Cultivation could fail in detecting pathogens that failed in competitive growth environments. It was interesting to see that many patients were infected by more than one pathogen, which might cause potential competition between different pathogens in CCT process (**S7 Table**). For example, *A. baumannii* was found to be the most competitive bacteria in cultivation, which may be due to its fastest growth rate. On the other hand, *P. aeruginosa* seemed to be relatively the weakest one among them, which was usually concealed in the cultivation with other species existing (sample A16, B19-3, B21-1, B21-2 after we excluded all samples with *A. baumannii* existing).

When evaluating the clinical benefit from SSBD, the quicker directed therapy adjustment for patients in the experiment group (Exp: 10.2±8.8 hours vs. Con: 96.0±35.1 hours, p<0.0001, Mann-Whitney test) could shorten the empirical anti-infection time and seemed to alleviate illness severity (APACHE II score) during the validation stage II with the help of the SSBD. Some clinical parameters were also showed good tendency in the experiment group patients on day 7, including the absolute value of white blood cell (WBC), the cases number with abnormal body temperature improved, and the clinical anti-infection efficiency (**S6 Table**). It implied that appropriate antibiotic treatment guided by in-time pathogenic information would alleviate acute physiological illness. Nevertheless, at the endpoint, clinical outcomes showed no differences between the two groups, which may due to the insufficient patient numbers.

Despite the size in our intervention stage, there were still some aspects that have not been considered. 1) Resistance genes were not included in the study. Multi-drug resistant organisms (MDROs) prevailed in ICU [24, 25], which might not improve the situation of patients even with accurate pathogenic information. There were a few cases (e.g. B07, B25 and B35 patients) showing no signs of clearing the bacterial infection. 2) The 10 designed pathogens were originated from sepsis, which might not completely overlap with pathogens of severe pneumonia, though pneumonia is one of the most common causes of sepsis. The panel pathogens could be optimized flexibly for meeting diverse clinical needs in the ICU. 3) Other potential pathogenic microbes, such as viruses and fungus, might affect the clinical outcomes considering the complexity of ICU patients.

Previous studies showed that polymicrobial pneumonia is related to an increased risk of inappropriate antimicrobial treatment [26]. In both phases, a total of 55 samples were identified as polymicrobial infections by SSBD, while only 14 samples were identified as polymicrobial infections by CCT, which suggested that SSBD could provide more precise pathogenic bacteria information than CCT, especially for those patients with polymicrobial infections. On the other hand, nosocomial infections contribute to a considerable proportion of deaths in ICU patients with severe pneumonia [23]. Although SSBD identified similar ratio of nosocomial infection events with CCT (**Fig 5G**), SSBD provided more timely information for clinical control and response, which might improve the clinical medication decision in ICU.

As anticipated, SSBD performed well with high sensitivity and specificity in rapid pathogens identification, and it reduced TTAT, which was associated with more rapid administration of appropriate antimicrobial therapy in the experiment cases. SSBD also has enormous potential in expanding pathogens from different diseases with much more pathogen genomes concluded. We believe that SSBD is competent in the role of clinical application and able to be applied in more clinical research.

## Data Availability

All data produced in the present work are contained in the manuscript.

## Figure Legends

**Supplementary Figure 1.**
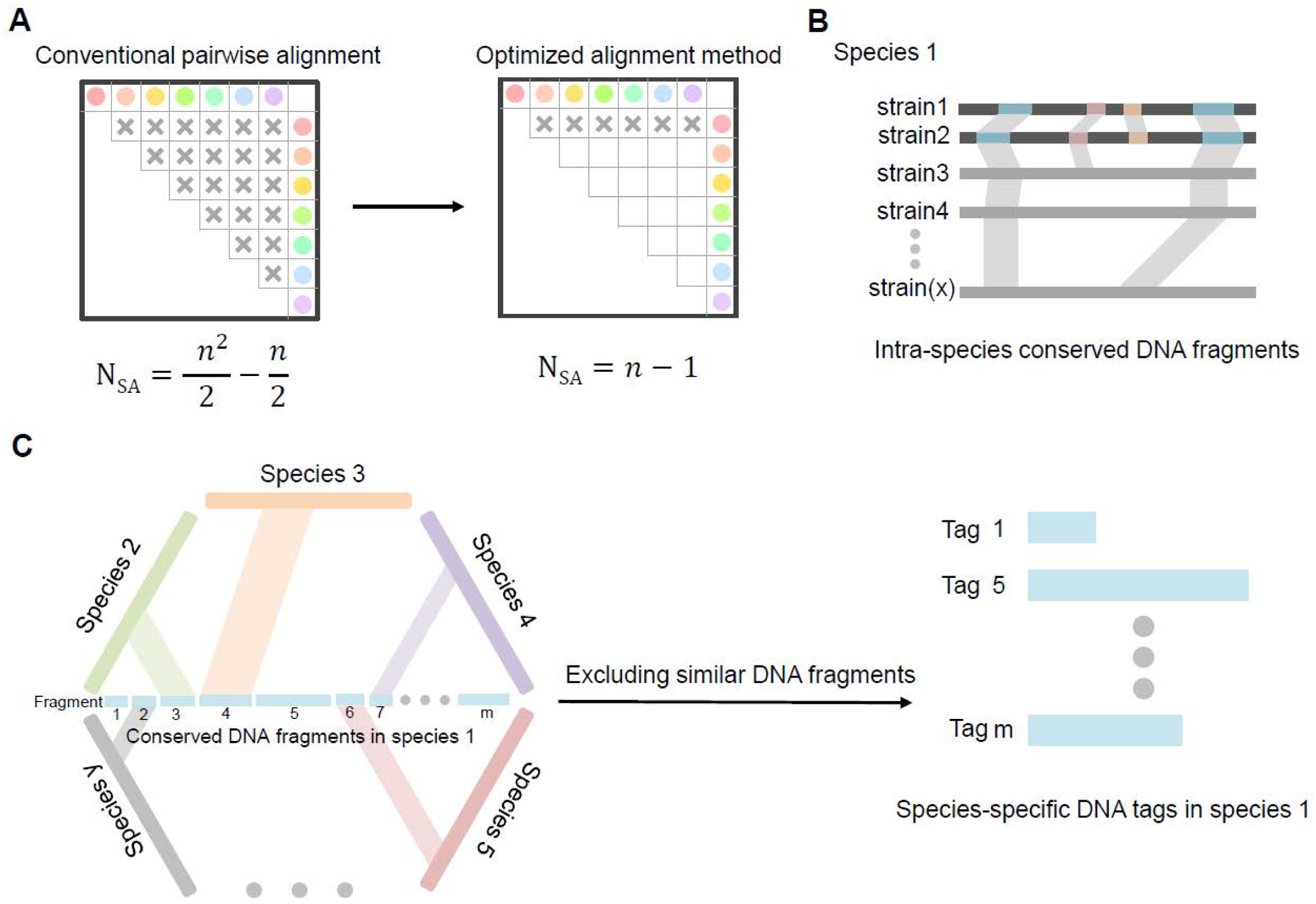

**Supplementary Figure 2.**
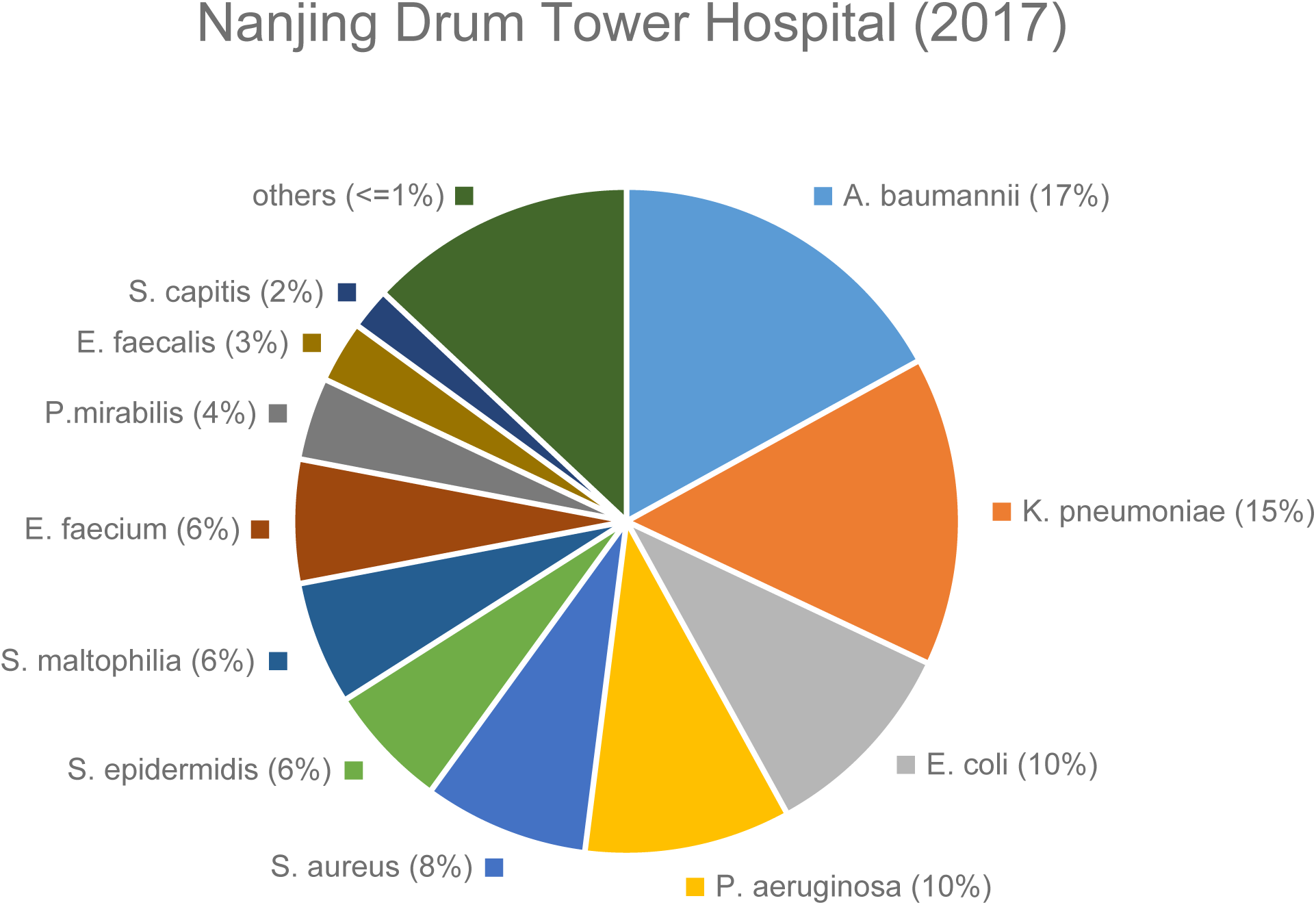

**Supplementary Figure 3.**
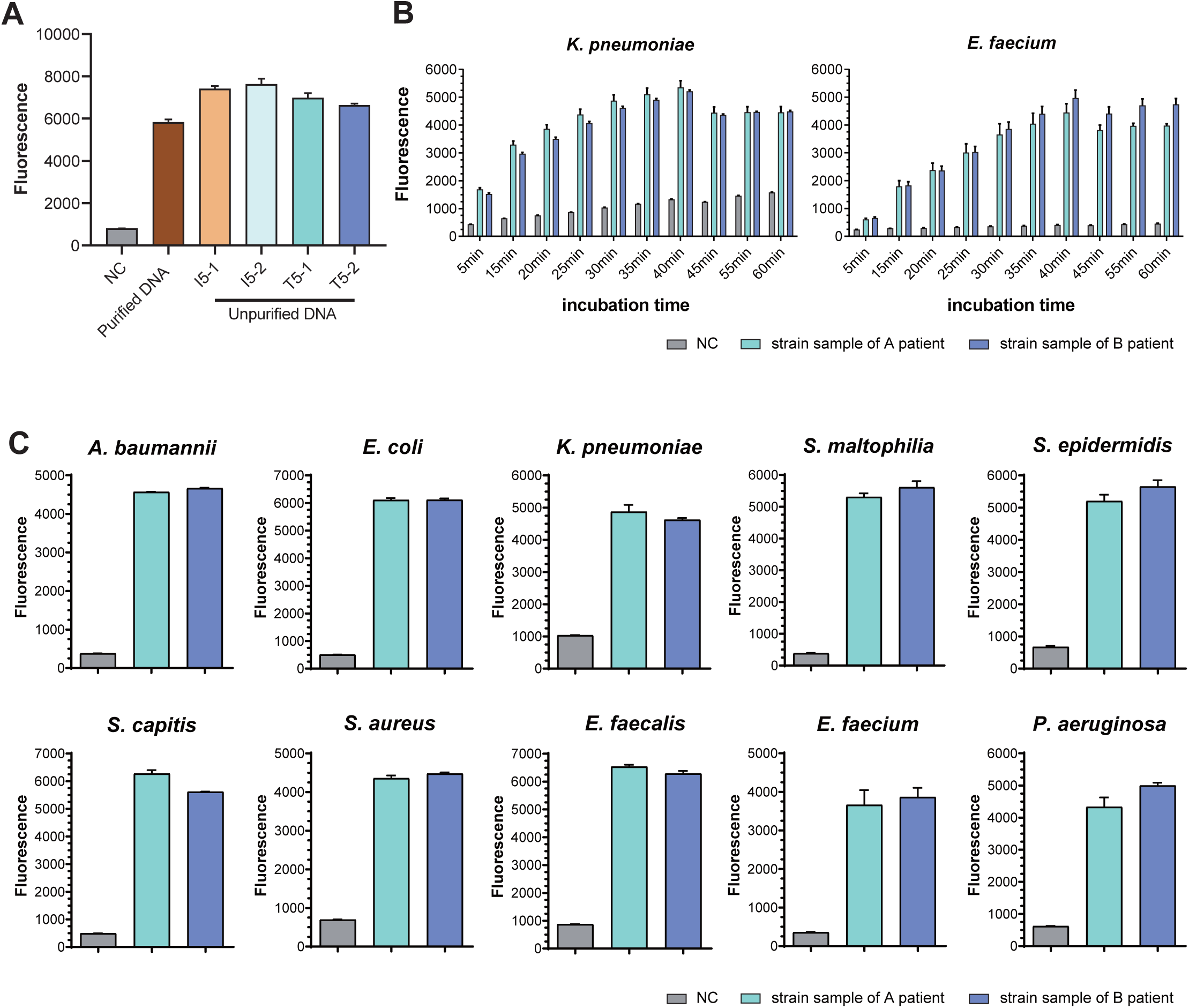

**Supplementary Figure 4.**
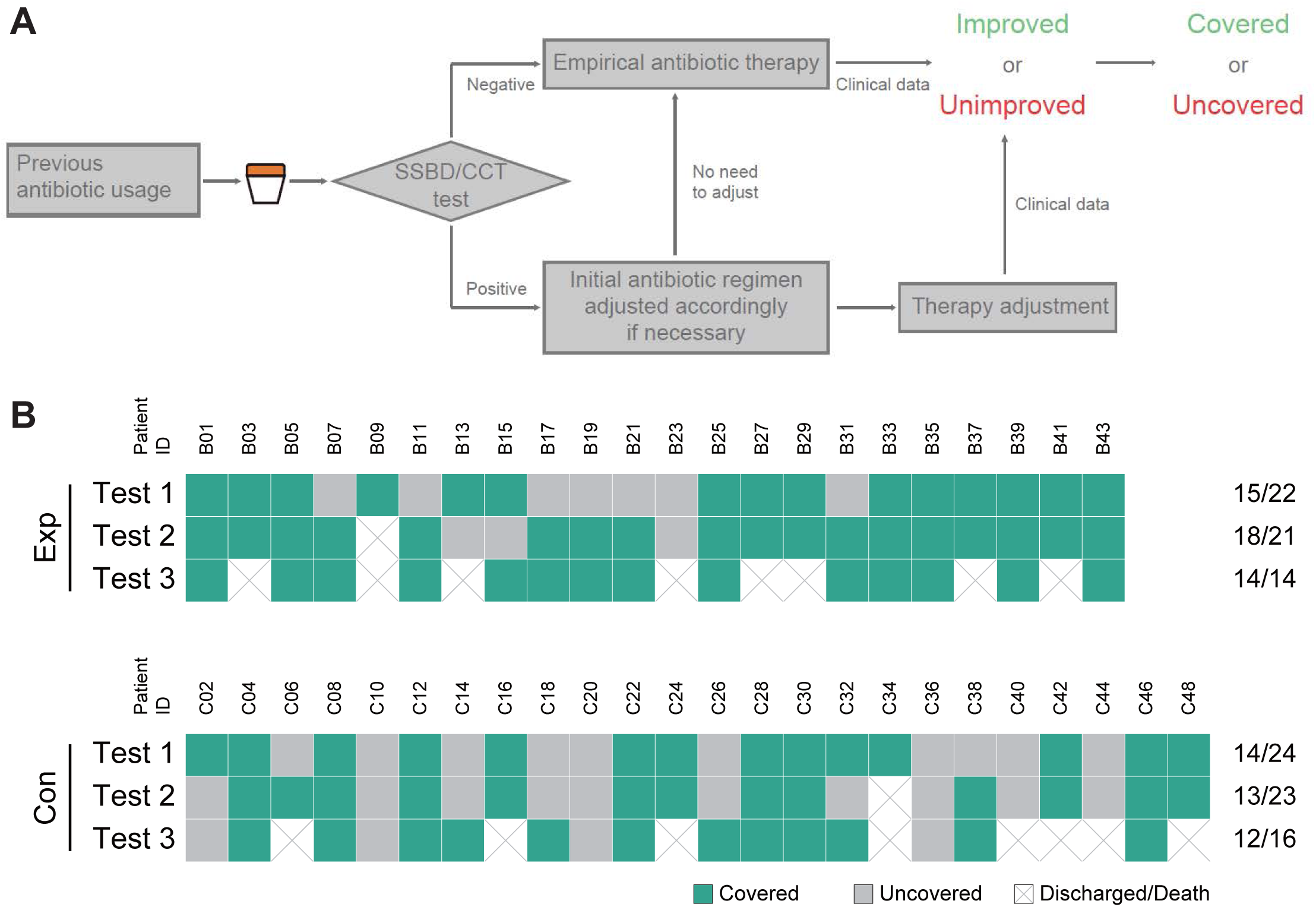

**Supplementary Figure 5.**
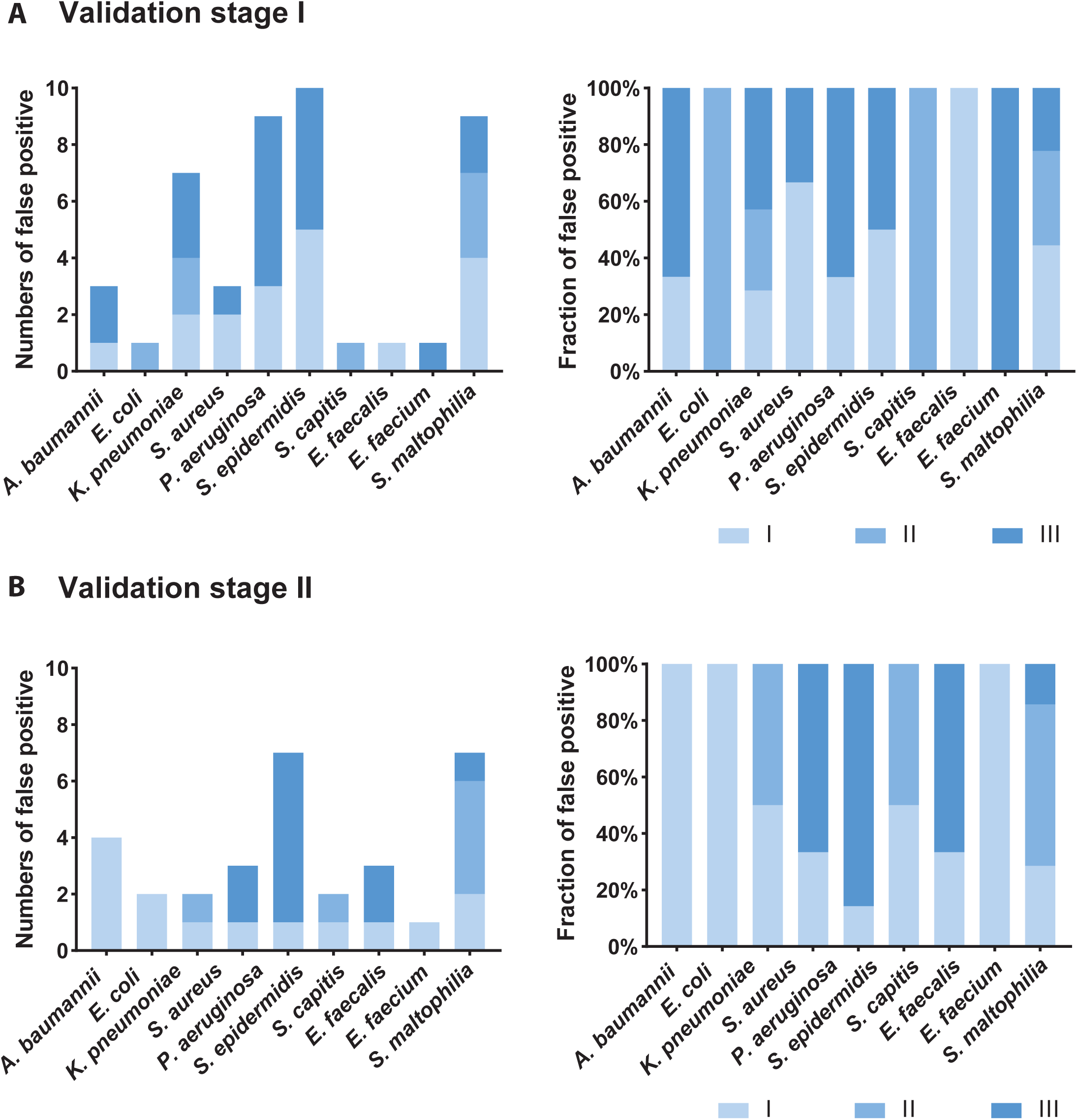

## References

1. Singer M, Deutschman CS, Seymour CW, Shankar-Hari M, Annane D, Bauer M, et al. The Third International Consensus Definitions for Sepsis and Septic Shock (Sepsis-3). Jama-Journal of the American Medical Association. 2016;315(8):801–10.

2. Ferrer R, Martinez ML, Goma G, Suarez D, Alvarez-Rocha L, de la Torre MV, et al. Improved empirical antibiotic treatment of sepsis after an educational intervention: the ABISS-Edusepsis study. Crit Care. 2018;22(1):167.

3. Kumar A, Roberts D, Wood KE, Light B, Parrillo JE, Sharma S, et al. Duration of hypotension before initiation of effective antimicrobial therapy is the critical determinant of survival in human septic shock. Critical Care Medicine. 2006;34(6):1589–96.

4. Pulia M, Redwood R. Empiric Antibiotic Prescribing for Suspected Sepsis: A Stewardship Balancing Act. Am J Med Sci. 2020;360(6):613–4.

5. Seymour CW, Gesten F, Prescott HC, Friedrich ME, Iwashyna TJ, Phillips GS, et al. Time to Treatment and Mortality during Mandated Emergency Care for Sepsis. New England Journal of Medicine. 2017;376(23):2235–44.

6. Abd El-Aziz NK, Gharib AA, Mohamed EAA, Hussein AH. Real-time PCR versus MALDI-TOF MS and culture-based techniques for diagnosis of bloodstream and pyogenic infections in humans and animals. J Appl Microbiol. 2021;130(5):1630–44.

7. Zhou J, Qian C, Zhao M, Yu X, Kang Y, Ma X, et al. Epidemiology and Outcome of Severe Sepsis and Septic Shock in Intensive Care Units in Mainland China. Plos One. 2014;9(9).

8. Chen H, Yin Y, Gao H, Guo Y, Dong Z, Wang X, et al. Clinical Utility of In-house Metagenomic Next-generation Sequencing for the Diagnosis of Lower Respiratory Tract Infections and Analysis of the Host Immune Response. Clinical Infectious Diseases. 2020;71:S416–S26.

9. Wang S, Ai J, Cui P, Zhu Y, Wu H, Zhang W. Diagnostic value and clinical application of next-generation sequencing for infections in immunosuppressed patients with corticosteroid therapy. Annals of Translational Medicine. 2020;8(5).

10. Edin A, Eilers H, Allard A. Evaluation of the Biofire Filmarray Pneumonia panel plus for lower respiratory tract infections. Infectious Diseases. 2020;52(7):479–88.

11. Jamal W, Al Roomi E, AbdulAziz LR, Rotimi VO. Evaluation of Curetis Unyvero, a Multiplex PCR-Based Testing System, for Rapid Detection of Bacteria and Antibiotic Resistance and Impact of the Assay on Management of Severe Nosocomial Pneumonia. Journal of Clinical Microbiology. 2014;52(7):2487–92.

12. Trotter AJ, Aydin A, Strinden MJ, O’Grady J. Recent and emerging technologies for the rapid diagnosis of infection and antimicrobial resistance. Current Opinion in Microbiology. 2019;51:39–45.

13. Nina Dombrowski, Tom A. Williams, Jiarui Sun, Benjamin J. Woodcroft, Jun-Hoe Lee, Bui Quang Minh, Christian Rinke & Anja Spang. Undinarchaeota illuminate DPANN phylogeny and the impact of gene transfer on archaeal evolution. Nature Communication. 2020. 11:3939.

14. Ilana Lauren Brito. Examining horizontal gene transfer in microbial communities. Nature Reviews Microbiology. 2021; 19(7): 442–453.

15. Mathieu Groussin, Mathilde Poyet, Elevated rates of horizontal gene transfer in the industrialized human microbiome. Cell. 2021; 184: 2053–2067.

16. C. Maslunka, V. Gurtler and R.J. Seviour. The impact of horizontal gene transfer on targeting the internal transcribed spacer region (ITS) to identify Acinetobacter junii strains. Journal of Applied Microbiology. 2015; 118: 1435–1443.

17. José M. Padial and Ignacio De la Riva. A paradigm shift in our view of species drives current trends in biological classification. Biol. Rev. 2021; 96:731–751.

18. De Pascale G, Bello G, Tumbarello M, Antonelli M: Severe pneumonia in intensive care: cause, diagnosis, treatment and management: a review of the literature. Current Opinion in Pulmonary Medicine 2012, 18(3):213–221.

19. Sakr Y, Jaschinski U, Wittebole X, Szakmany T, Lipman J, Namendys-Silva SA, Martin-Loeches I, Leone M, Lupu M-N, Vincent J-L et al: Sepsis in Intensive Care Unit Patients: Worldwide Data From the Intensive Care over Nations Audit. Open Forum Infectious Diseases 2018, 5(12).

20. Janice S. Chen, Enbo Ma, Lucas B. Harrington, Maria Da Costa, Xinran Tian, Joel M. Palefsky, Jennifer A. Doudna. CRISPR-Cas12a target binding unleashes indiscriminate single-stranded DNase activity. Science. 2018;360:436–439.

21. Jonathan S. Gootenberg, Omar O. Abudayyeh, Max J. Kellner, Julia Joung, James J. Collins, Feng Zhang. Multiplexed and portable nucleic acid detection platform with Cas13, Cas12a, and Csm6. Science. 2018;360:439–444.

22. Andreia S. Azevedo, Carina Almeida, Luís F. Melo & Nuno F. Azevedo. Impact of polymicrobial biofilms in catheterassociated urinary tract infections. Crit Rev Microbiol. 2016. 43(4):423–439.

23. Rafael Zaragoza, Pablo Vidal-Cortés, Gerardo Aguilar, Marcio Borges, Emili Diaz, Ricard Ferrer, Emilio Maseda, Mercedes Nieto, Francisco Xavier Nuvials, Paula Ramirez, Alejandro Rodriguez, Cruz Soriano, Javier Veganzones and Ignacio Martín-Loeches. Update of the treatment of nosocomial pneumonia in the ICU. 2020. 24(1):383.

24. Liu J, Zhang L, Pan J, Huang M, Li Y, Zhang H, et al. Risk Factors and Molecular Epidemiology of Complicated Intra-Abdominal Infections With Carbapenem-Resistant Enterobacteriaceae: A Multicenter Study in China. J Infect Dis. 2020;221(Suppl 2):S156–S63.

25. Yang Q, Zhang H, Yu Y, Kong H, Duan Q, Wang Y, et al. In Vitro Activity of Imipenem/Relebactam Against Enterobacteriaceae Isolates Obtained from Intra-abdominal, Respiratory Tract, and Urinary Tract Infections in China: Study for Monitoring Antimicrobial Resistance Trends (SMART), 2015-2018. Clin Infect Dis. 2020;71(Suppl 4):S427–S35.

26. Lisa Karner, Susanne Drechsler, Magdalena Metzger, Ara Hacobian, Barbara Schädl, Paul Slezak, Johannes Grillari and Peter Dungel. Antimicrobial photodynamic therapy fighting polymicrobial infections – a journey from in vitro to in vivo. Photochemical & Photobiological Sciences. 2020. 19(10):1332–1343.

27. Carolyn B Ibberson and Marvin Whiteley. The social life of microbes in chronic infection. Current Opinion in Microbiology. 2020. 53:44–50.

