## Supplementary methods and tables for "Species-specific bacterial detector for fast pathogen diagnosis of severe pneumonia patients in the intensive care unit"

**Supplementary Materials**

**Principles of screening species-specific DNA-tags**

Our core principles for screening species-specific DNA fragments were as follows: 1) Multiple isolated strains from the same bacterial species were included to ensure the intra-species conservation of selected DNA fragments; 2) Those similar DNA fragments of intra-species conserved fragments in genomes from other bacteria were excluded to ensure inter-species specificity. We developed an original workflow and optimized the algorithm to more efficiently achieve our purposes compared with conventional pairwise alignment (**S1A Fig**). Firstly, conserved DNA regions were obtained by aligning the genomes of two strains within a given species, downsizing the genome to the regional scale. Those conserved DNA regions were performed alignments with the genomes of other strains from the same species, only shared DNA regions in all the strains are retained, downsizing conserved genomic regions to intra-species conserved DNA fragments (**S1B Fig**). Secondly, to achieve inter-species specificity of DNA tags, intra-species conserved DNA fragments were performed alignments with the genomes of other bacterial species. Those similar fragments of intra-species conserved DNA fragments in genomes from other bacteria species were excluded. After two-step screening, we finally obtained species-specific DNA tags (**S1C Fig**).

**Extracting samples**

Extracting bronchoalveolar lavage fluid (BALF) for the enrolled patients were under mild anesthesia via tracheal intubation or tracheotomy entering the infected bronchus. 30 mL saline was injected into batches quickly and recollected by using negative pressure lower than 13.3 kPa. In the validation stage I, 5 mL collected samples were cultivated as usual, and another 5 mL samples were sent for SSBD testing.

**Reaction process**

Briefly, DNA of BALF samples was extracted using Quick–DNA/RNA™ Pathogen Miniprep Kit (Zymo Research) according to the manual and diluted to 100 ng/μL. The DNA samples were then amplified using designed specific primers for 10 bacteria. Then, PCR products, Cas12a-crRNA complexes and the reporter DNA probe were added to the reaction system. Finally, the fluorescence signals were detected after incubation for 30 mins at 37 ℃. Some samples were sent for next-generation sequencing (IngeniGen XunMinKang Biotechnology Inc. Hangzhou, Zhejiang, China).

**Clinical operation process**

In the validation stage II, BALFs were collected from each patient on the first day, day 3–5, and day 7+ unless the airway was removed. All samples were sent for microgram cultivation, and simultaneously the samples taken from the experiment group were tested by SSBD. Once BALF results were available, at least two clinical experts (antimicrobial stewardship) discussed and decided on antibiotics adjustment according to the results and other clinical data. Additionally, the experts would assess each patient's receiving antibiotics coverage rate at different times and other clinical outcomes retrospectively.

Treatment and outcomes data, such as evaluation of therapeutic effectiveness at day 3, 7, 10 and 14, time of mechanical ventilation and vasopressor support from enrollment to day 28, occurrence of antibiotic-associated diarrhea, and mortality on day 28 were also recorded and analyzed. Evaluation of therapeutic effectiveness was conducted by two senior clinicians according to these parameters.: (1) whether fever and purulent secretion were improved or not; (2) whether leukocytosis or leukopenia got better or not; (3) whether radiological pulmonary infiltrate absorbed partly or not; (4) whether oxygenation index was improved or not; (5) whether hemodynamic instability was rectified gradually or not.

**SSBD diagnostic report**

For every single experiment, we test the fluorescence value of clinical separated positive bacteria strains as positive control (PC) and deionized water as negative control (NC). For each experiment, we test the fluorescence value of BALF (shown as F) with a microplate reader. When F/NC > 2, it is the signal that bacteria detected by our method (Supplementary Figure 1A after PCR, F showing significance difference from the NC when F approximately 2 times of the NC). We use I (interval) as our point of distinction. I= (PC - 2NC) / 3. When 2NC < F < 2NC + I, the bacteria strength level is defined as level I. When 2NC + I < F < 2NC + 2I, the bacteria strength level is defined as level II. When F > 2NC + 2I, the bacteria strength level is defined as level III. Different levels of bacterial strength can roughly represent different bacterial copies according to our method lowest detection rate (Level I: 10^-15^ M–10^-14^ M, Level II: 10^-14^ M–10^-13^ M, Level III: over 10^-13^ M, Fig 3B). We separated different testing values as different levels, suggesting that our method could test pathogen strength to some extent and give pathogens strength for drug usage.

**Evaluation of antibiotics coverage**

For three individually designated BALF tests, the rate of antibiotics coverage was calculated in all groups. We calculated coverage rate from two aspects. For each test, we calculated rate of covered samples among all samples tested. For each patient, we calculated their coverage rate by counting covered test numbers within all tests that had been taken. Evaluation of antibiotics coverage was made by two experts retrospectively, according to microbial, antimicrobial susceptibility tests (AST) and clinical treatment effect. The details were as follow:

- Our test result is negative (from the experiment group), or microbial cultivation is negative (from the control group). If it is deemed effective on the clinical signs (1), it is judged as antibiotics covering. If it is deemed clinically invalid, it is judged as antibiotics uncovering.
- Our test result is positive (from the experiment group), or microbial cultivation is positive (from the control group). If AST (from experiment group or control group) is shown sensitive to the antibiotics, no matter the clinical signs are effective or not, it is judged as antibiotics covering. If AST is shown resistant to using antibiotics, whether antibiotics covering or not is judged by clinical effectiveness. It is identified as covered when clinically effective and uncovered when clinically invasive.

When BALF is not collected for a test or microbial cultivation from the day on, whether with covering antibiotics or not on the day is judged on clinical effectiveness.

**Supplementary figure legends**

**Supplementary Figure 1. Diagram of core principles for** **screening species-specific DNA-tags.**

A. Optimizing the algorithm of sequence alignment. Abbreviations: SA, sequence alignment; N, number of double sequence alignment; n, number of sequences.

B. Schematic map of screening intra-species conserved DNA fragments.

C. Schematic map of screening species-specific DNA tags.

**Supplementary Figure 2. Epidemic data of pathogens in the drum tower hospital ICU in 2017.**

**Supplementary Figure 3. SSBD development and effectiveness validation.**

A. SSBD results of purified and unpurified DNA.

B. SSBD results of reaction time gradient with Cas12a.

Fluorescence values of *K. pneumoniae* and *E. faecium* by Cas12a through different incubation times after PCR. Gray represented NC, namely the fluorescence values of PCR products of using DEPC-H2O as input. Green and blue represented the fluorescence values of bacteria strains from different patients. Each group had three repeats. Error bars indicated mean ± SEM of fluorescence value.

C. SSBD results of 10 pathogenic bacteria with Cas12a.

Gray represented NC, namely the fluorescence values of PCR products of using DEPC-H2O as input. Green and blue represented the fluorescence values of bacteria strains from different patients. Each group had three repeats. Error bars indicated mean ± SEM of fluorescence value. All p < 0.001 with unpaired t-test.

**Supplementary Figure 4. Judgment process and results of antibiotics coverage.**

A. Judgment process of antibiotics coverage.

B. The raw results of antibiotics coverage in two groups. Exp meant the experimental group, and Con meant the control group.

**Supplementary Figure 5. Analysis of false-positive samples.**

Numbers and fractions of different strength levels among all false-positive samples of each bacteria species in the validation stage I (A) and II (B). Strength could be seen roughly as bacterial amounts (level I-level III, the definition could be seen in the Supplementary Material). False-positive situations meant pathogenic bacteria detected by SSBD but not by CCT in a given BALF sample.

**Abbreviations:**

*Acinetobacter baumannii*: *A. baumannii*

*Enterococcus faecalis*: *E. faecalis*

*Enterococcus faecium*: *E. faecium*

*Escherichia coli*: *E. coli*

*Klebsiella pneumoniae*: *K. pneumoniae*

*Pseudomonas aeruginosa*: *P. aeruginosa*

*Staphylococcus aureus*: *S. aureus*

*Staphylococcus capitis*: *S. capitis*

*Staphylococcus epidermidis*: *S. epidermidis*

*Stenotrophomonas maltophilia*: *S. maltophilia*

N: Negative

**Supplementary Table 1: Primers used in experiments.**

| **Name** | **Sequence (5' -> 3')** | **Target** | **Product length (bp)** |
| --- | --- | --- | --- |
| pGL3-amplify-F | GAAGATGGAACCGCTGGAGA | pGL3 | 597 |
| pGL3-amplify-R | GCAGGCAGTTCTATGAGGCA |  |  |
| Aba-amplify-F | CACAGCGTTTAACCCATGCC | *A. baumannii* | 564 |
| Aba-amplify-R | TATCGCCACCTGCACAGAAG |  |  |
| Eco-amplify-F | GTTCCTGACTATCTGGCGGG | *E. coli* | 371 |
| Eco-amplify-R | GCTTCCTGACTCCAGACACC |  |  |
| Kpn-amplify-F | CATGGGCATATCGACGGTCA | *K. pneumoniae* | 740 |
| Kpn-amplify-R | CCTGCAACATAGGCCAGTGA |  |  |
| Sau-amplify-F | AGGTGCAGTAGACGCATAGC | *S. aureus* | 563 |
| Sau-amplify-R | CATTCGCTGCGCCAATACAA |  |  |
| Pae-amplify-F | TCTCTCTATCACGCCGGTCA | *P. aeruginosa* | 467 |
| Pae-amplify-R | TCGCATCGAGGTATTCCAGC |  |  |
| Sep-amplify-F | CACGCATGGCACTAGGTACA | *S. epidermidis* | 383 |
| Sep-amplify-R | CGAAAAAGAGTTGTCCTTGTTGA |  |  |
| Sca-amplify-F | GGTTCAGTCATCCCCACGTT | *S. capitis* | 591 |
| Sca-amplify-R | CAGCTGCGACAACTGCTTAC |  |  |
| Efa-amplify-F | CGGCAAGTTTGGAAGCAGAC | *E. faecalis* | 627 |
| Efa-amplify-R | CAGCGCCTAGTCCTTGTGAT |  |  |
| Efm-amplify-F | ATCGGAAATCGGTGTGGCTT | *E. faecium* | 507 |
| Efm-amplify-R | TCAAATGCATCCCTGTGCCT |  |  |
| Sma-amplify-F | CGCCTCCCGTTTACAGATTA | *S. maltophilia* | 356 |
| Sma-amplify-R | TCGGCTCCACCACATACAC |  |  |

**Supplementary Table 2: Oligonucleotide templates for synthesis of crRNAs.**

| **Name** | **Sequence (5′->3′)** | **Target** |
| --- | --- | --- |
| T7-Forward | TAATACGACTCACTATAGGT |  |
| LbcrRNA-Reverse-PGL3 | ATGTGACGAACGTGTACATCGACTATCTACACTTAGTAGAAATTACCTATAGTGAGTCGTATTA | PGL3 |
| LbcrRNA-Reverse-Aba | TTCAAGTAATTCTTCTTTACATCTACACTTAGTAGAAATTACCTATAGTGAGTCGTATTA | *A. baumannii* |
| LbcrRNA-Reverse-Eco | CTTGCCATCATAGCGACCGTATCTACACTTAGTAGAAATTACCTATAGTGAGTCGTATTA | *E. coli* |
| LbcrRNA-Reverse-Kpn | AAGATGGCGATTACCGCAGTATCTACACTTAGTAGAAATTACCTATAGTGAGTCGTATTA | *K. pneumoniae* |
| LbcrRNA-Reverse-Sau | CCTCAGCAAGTTCACGTTGTATCTACACTTAGTAGAAATTACCTATAGTGAGTCGTATTA | *S. aureus* |
| LbcrRNA-Reverse-Pae | CTCCTCATGTGTGTTTACAAATCTACACTTAGTAGAAATTACCTATAGTGAGTCGTATTA | *P. aeruginosa* |
| LbcrRNA-Reverse-Sep | TCTCTAATTGATGAATATTAATCTACACTTAGTAGAAATTACCTATAGTGAGTCGTATTA | *S. epidermidis* |
| LbcrRNA-Reverse-Sca | TATTGATTAATAAGGTGATTATCTACACTTAGTAGAAATTACCTATAGTGAGTCGTATTA | *S. capitis* |
| LbcrRNA-Reverse-Efa | TCAGCTGTGTTATTTGGTGCATCTACACTTAGTAGAAATTACCTATAGTGAGTCGTATTA | *E. faecalis* |
| LbcrRNA-Reverse-Efm | TGTATATAAGTTCAGGTAGTATCTACACTTAGTAGAAATTACCTATAGTGAGTCGTATTA | *E. faecium* |
| LbcrRNA-Reverse-Sma | CCTGGTCGCAGGTGTCATGCATCTACACTTAGTAGAAATTACCTATAGTGAGTCGTATTA | *S. maltophilia* |

**Supplementary Table 3: SSBD and CCT results of BALF samples in the validation stage I.**

| **Sample ID** | **SSBD Results** | **CCT Results** | **NGS Results** |
| --- | --- | --- | --- |
| A01 | *S. aureus* (I) | N | *S. aureus* |
| A02 | *A. baumannii* (III) | *A. baumannii* |  |
| A03 | *A. baumannii* (III)  *S. aureus* (I) | *A. baumannii*  *S. aureus* | *A. baumannii*  *S. aureus* |
| A04 | *A. baumannii* (III)  *K. pneumoniae* (II)  *S. maltophilia* (II) | N |  |
| A05 | *A. baumannii* (III)  *K. pneumoniae* (III)  *P. aeruginosa* (III) | *K. pneumoniae*  *P. aeruginosa* |  |
| A06 | *A. baumannii* (III) | *A. baumannii* |  |
| A07 | *S. maltophilia* (II) | *S. maltophilia* |  |
| A08 | *A. baumannii* (III) | *A. baumannii* |  |
| A09 | *A. baumannii* (II)  *S. aureus* (I) | *A. baumannii* |  |
| A10 | *A. baumannii* (III)  *K. pneumoniae* (III)  *P. aeruginosa* (I) | *A. baumannii*  *K. pneumoniae* |  |
| A11 | *A. baumannii* (III)  *K. pneumoniae* (I) | *A. baumannii* |  |
| A12 | N | N |  |
| A13 | *A. baumannii* (II) | *A. baumannii* |  |
| A14 | N | N |  |
| A15 | *K. pneumoniae* (III) | *K. pneumoniae* |  |
| A16 | *K. pneumoniae* (III)  *P. aeruginosa* (III) | *K. pneumoniae* |  |
| A17 | N | N |  |
| A18 | *K. pneumoniae* (I)  *P. aeruginosa* (III) | *K. pneumoniae*  *P. aeruginosa* |  |
| A19 | *P. aeruginosa* (I) | *P. aeruginosa* |  |
| A20 | *E. faecium* (III)  *S. capitis* (II) | N | *E. faecium*  *S. capitis* |
| A21 | *A. baumannii* (III)  *K. pneumoniae* (III)  *S. aureus* (I) | *A. baumannii*  *S. aureus* |  |
| A22 | *A. baumannii* (III)  *S. epidermidis* (III) | *A. baumannii* |  |
| A23 | N | N |  |
| A24 | *A. baumannii* (III) | *A. baumannii* |  |
| A25 | N | N | N |
| A26 | *P. aeruginosa* (III) | *P. aeruginosa* |  |
| A27 | *A. baumannii* (III)  *S. epidermidis* (III) | *A. baumannii* | *A. baumannii*  *S. epidermidis* |
| A28 | *A. baumannii* (II)  *P. aeruginosa* (III) | *A. baumannii* |  |
| A29 | *S. epidermidis* (I)  *S. maltophilia* (I) | N |  |
| A30 | *A. baumannii* (III)  *S. aureus* (III)  *S. epidermidis* (I) | *A. baumannii* |  |
| A31 | *K. pneumoniae* (I)  *P. aeruginosa* (I)  *S. epidermidis* (I) | N | *K. pneumoniae* |
| A32 | *P. aeruginosa* (I)  *S. epidermidis* (I) | N |  |
| A33 | *S. aureus* (I)  *S. maltophilia* (II) | *S. aureus* |  |
| A34 | N | N | *A. baumannii* |
| A35 | *A. baumannii* (III) | *A. baumannii* |  |
| A36 | *A. baumannii* (III)  *S. maltophilia* (I) | *A. baumannii* |  |
| A37 | *A. baumannii* (III)  *S. epidermidis* (III) | *A. baumannii* |  |
| A38 | *A. baumannii* (III) | *A. baumannii* |  |
| A39 | *A. baumannii* (III) | *A. baumannii* |  |
| A40 | N | N |  |
| A41 | N | N |  |
| A42 | *A. baumannii* (I) | N |  |
| A43 | *A. baumannii* (III) | *A. baumannii* |  |
| A44 | *A. baumannii* (III) | *A. baumannii* | *A. baumannii* |
| A45 | *A. baumannii* (III)  *K. pneumoniae* (II) | *A. baumannii* |  |
| A46 | *A. baumannii* (III) | *A. baumannii* |  |
| A47 | *A. baumannii* (I) | *A. baumannii* |  |
| A48 | *A. baumannii* (III) | *A. baumannii* |  |
| A49 | *A. baumannii* (III)  *S. aureus* (III)  *S. maltophilia* (III) | *A. baumannii*  *S. aureus* | *A. baumannii*  *S. aureus*  *S. maltophilia* |
| A50 | *S. epidermidis* (III) | N |  |
| A51 | N | N |  |
| A52 | N | N |  |
| A53 | *K. pneumoniae* (III) | *K. pneumoniae* |  |
| A54 | N | N |  |
| A55 | *A. baumannii* (III)  *E. coli* (II)  *K. pneumoniae* (III)  *S. maltophilia* (III) | *A. baumannii* |  |
| A56 | *S. epidermidis* (III)  *E. faecalis* (I) | N |  |
| A57 | N | N |  |
| A58 | *P. aeruginosa* (III) | N |  |
| A59 | *S. maltophilia* (I) | *S. maltophilia* | *S. aureus*  *S. maltophilia* |
| A60 | *A. baumannii* (I)  *P. aeruginosa* (III) | *A. baumannii*  *P. aeruginosa* |  |
| A61 | *A. baumannii* (II) | *A. baumannii* |  |
| A62 | *K. pneumoniae* (II)  *S. maltophilia* (III) | *K. pneumoniae*  *S. maltophilia* |  |
| A63 | *A. baumannii* (III) | *A. baumannii* |  |
| A64 | *A. baumannii* (III) | *A. baumannii* |  |
| A65 | *A. baumannii* (III)  *P. aeruginosa* (I)  *S. maltophilia* (II) | *A. baumannii*  *P. aeruginosa* |  |
| A66 | *K. pneumoniae* (II) | *K. pneumoniae* |  |
| A67 | *S. aureus* (III) | *S. aureus* |  |
| A68 | *A. baumannii* (III)  *P. aeruginosa* (III) | *A. baumannii* |  |
| A69 | *A. baumannii* (I)  *P. aeruginosa* (III) | *A. baumannii* | *P. aeruginosa* |
| A70 | *A. baumannii* (I) | *A. baumannii* |  |
| A71 | N | N |  |
| A72 | *A. baumannii* (III)  *S. maltophilia* (I) | *A. baumannii* |  |
| A73 | *A. baumannii* (III)  *K. pneumoniae* (III)  *P. aeruginosa* (III) | *A. baumannii*  *P. aeruginosa* |  |
| A74 | *A. baumannii* (III) | *A. baumannii* |  |
| A75 | *A. baumannii* (III)  *P. aeruginosa* (III) | *A. baumannii* |  |
| A76 | *A. baumannii* (I)  *P. aeruginosa* (III)  *S. maltophilia* (I) | *P. aeruginosa* |  |
| A77 | *S. epidermidis* (I) | N |  |

**Supplementary Table 4: SSBD and CCT results of BALF samples from experiment group (n=22) and control group (n=24) during the validation stage II.**

| **Patient ID** | **Sample ID** | **Test No.** | **SSBD Results** | **CCT Results** | **NGS Results** |
| --- | --- | --- | --- | --- | --- |
| **Exp.** |  |  |  |  |  |
| B01 | B01-1 | Test 1 | N | N |  |
|  | B01-2 | Test 2 | N | N | N |
|  | B01-3 | Test 3 | *A. baumannii* (III) | *A. baumannii* |  |
| B03 | B03-1 | Test 1 | N | N | N |
|  | B03-2 | Test 2 | *A. baumannii* (III) | *A. baumannii* |  |
| B05 | B05-1 | Test 1 | *A. baumannii* (III)  *S. maltophilia* (II) | *A. baumannii* | *A. baumannii*  *S. maltophilia* |
|  | B05-2 | Test 2 | *A. baumannii* (III)  *S. maltophilia* (I) | *A. baumannii* |  |
|  | B05-3 | Test 3 | *A. baumannii* (III)  *S. maltophilia* (I) |  |  |
| B07 | B07-1 | Test 1 | *P. aeruginosa* (III) | *P. aeruginosa* | *P. aeruginosa* |
|  | B07-2 | Test 2 | *P. aeruginosa* (III) | *P. aeruginosa* |  |
|  | B07-3 | Test 3 | *P. aeruginosa* (III) | *P. aeruginosa* |  |
| B09 | B09-1 | Test 1 | *A. baumannii* (III)  *S. aureus* (III)  *S. capitis* (III)  *S. maltophilia* (III) | *A. baumannii* |  |
| B11 | B11-1 | Test 1 | *A. baumannii* (III)  *P. aeruginosa* (III)  *S. maltophilia* (II) | *A. baumannii* | *A. baumannii*  *P. aeruginosa*  *S. maltophilia* |
|  | B11-2 | Test 2 | *A. baumannii* (III)  *S. aureus* (I)  *S. maltophilia* (III) |  |  |
|  | B11-3 | Test 3 | *P. aeruginosa* (III) | N |  |
| B13 | B13-1 | Test 1 | *S. epidermidis* (I) | N | N |
|  | B13-2 | Test 2 | *K. pneumoniae* (I) | *K. pneumoniae* |  |
| B15 | B15-1 | Test 1 | *A. baumannii* (I)  *E. coli* (I)  *K. pneumoniae* (II) | N |  |
|  | B15-2 | Test 2 | *K. pneumoniae* (I)  *E. faecium* (I) |  |  |
|  | B15-3 | Test 3 | *E. faecium* (III) |  |  |
| B17 | B17-1 | Test 1 | *A. baumannii* (III) |  |  |
|  | B17-2 | Test 2 | *A. baumannii* (III) *K. pneumoniae* (III) |  |  |
|  | B17-3 | Test 3 | *A. baumannii* (I)  *K. pneumoniae* (I)  *E. faecalis* (I) | *A. baumannii* |  |
| B19 | B19-1 | Test 1 | *A. baumannii* (III)  *P. aeruginosa* (I)  *S. maltophilia* (II) | *A. baumannii* |  |
|  | B19-2 | Test 2 | *A. baumannii* (III)  *P. aeruginosa* (III)  *S. maltophilia* (III) | *A. baumannii*  *S. maltophilia* |  |
|  | B19-3 | Test 3 | *K. pneumoniae* (III)  *P. aeruginosa* (III)  *S. maltophilia* (III) | *K. pneumoniae*  *S. maltophilia* |  |
| B21 | B21-1 | Test 1 | *P. aeruginosa* (III)  *S. aureus* (III)  *S. maltophilia* (III) | *S. aureus*  *S. maltophilia* |  |
|  | B21-2 | Test 2 | *P. aeruginosa* (III)  *S. aureus* (III)  *S. maltophilia* (III) | *S. maltophilia* |  |
|  | B21-3 | Test 3 | *S. aureus* (I)  *S. maltophilia* (II) | N |  |
| B23 | B23-1 | Test 1 | *S. capitis* (I)  *S. maltophilia* (III) | *S. maltophilia* |  |
|  | B23-2 | Test 2 | *S. maltophilia* (III) | *S. maltophilia* |  |
| B25 | B25-1 | Test 1 | *A. baumannii* (II) | *A. baumannii* |  |
|  | B25-2 | Test 2 | *A. baumannii* (III) |  |  |
|  | B25-3 | Test 3 | *A. baumannii* (III) | *A. baumannii* |  |
| B27 | B27-1 | Test 1 | *S. maltophilia* (I) | N | N |
|  | B27-2 | Test 2 | *A. baumannii* (III) |  |  |
| B29 | B29-1 | Test 1 | *K. pneumoniae* (III) | *K. pneumoniae* | *K. pneumoniae* |
|  | B29-2 | Test 2 | *K. pneumoniae* (III) |  |  |
| B31 | B31-1 | Test 1 | *A. baumannii* (III)  *S. capitis* (III) | *A. baumannii* | *A. baumannii*  *S. capitis* |
|  | B31-2 | Test 2 | *A. baumannii* (III) |  |  |
|  | B31-3 | Test 3 | *A. baumannii* (I) | N |  |
| B33 | B33-1 | Test 1 | N | N |  |
|  | B33-2 | Test 2 | *A. baumannii* (I)  *S. epidermidis* (II) | N |  |
|  | B33-3 | Test 3 | N | N |  |
| B35 | B35-1 | Test 1 | *A. baumannii* (III) | *A. baumannii* | *A. baumannii* |
|  | B35-2 | Test 2 | *A. baumannii* (III) | *A. baumannii* |  |
|  | B35-3 | Test 3 | *A. baumannii* (III) | *A. baumannii* |  |
| B37 | B37-1 | Test 1 | N | N | *A. baumannii* |
|  | B37-2 | Test 2 | *P. aeruginosa* (I) |  |  |
| B39 | B39-1 | Test 1 | N | *P. aeruginosa* |  |
|  | B39-2 | Test 2 | *A. baumannii* (I)  *E. coli* (I)  *K. pneumoniae* (III)  *P. aeruginosa* (III) | *K. pneumoniae*  *P. aeruginosa* |  |
|  | B39-3 | Test 3 | *K. pneumoniae* (I)  *P. aeruginosa* (III) |  |  |
| B41 | B41-1 | Test 1 | N | N | N |
|  | B41-2 | Test 2 | N |  |  |
| B43 | B43-1 | Test 1 | N | N |  |
|  | B43-2 | Test 2 | *A. baumannii* (I)  *S. epidermidis* (I) |  |  |
|  | B43-3 | Test 3 | *A. baumannii* (II) | *A. baumannii* |  |
| **Con.** |  |  |  |  |  |
| C02 |  | Test 1 |  | *A. baumannii* |  |
|  |  | Test 2 |  | *A. baumannii* |  |
|  |  | Test 3 |  | *A. baumannii* |  |
| C04 |  | Test 1 |  | N |  |
|  |  | Test 2 |  | N |  |
|  |  | Test 3 |  | N |  |
| C06 |  | Test 1 |  | N |  |
|  |  | Test 2 |  | *A. baumannii* |  |
| C08 |  | Test 1 |  | *E. coli* |  |
|  |  | Test 2 |  | *E. coli* |  |
|  |  | Test 3 |  | *K. pneumoniae* |  |
| C10 |  | Test 1 |  | N |  |
|  |  | Test 2 |  | N |  |
|  |  | Test 3 |  | *A. baumannii* |  |
| C12 |  | Test 1 |  | N |  |
|  |  | Test 2 |  | *A. baumannii* |  |
|  |  | Test 3 |  | *A. baumannii* |  |
| C14 |  | Test 1 |  | *A. baumannii* |  |
|  |  | Test 2 |  | *A. baumannii* |  |
|  |  | Test 3 |  | *A. baumannii* |  |
| C16 |  | Test 1 |  | N |  |
|  |  | Test 2 |  | N |  |
| C18 |  | Test 1 |  | *A. baumannii* |  |
|  |  | Test 2 |  | *A. baumannii* |  |
|  |  | Test 3 |  | *A. baumannii* |  |
| C20 |  | Test 1 |  | N |  |
|  |  | Test 2 |  | *A. baumannii* |  |
|  |  | Test 3 |  | *A. baumannii* |  |
| C22 |  | Test 1 |  | N |  |
|  |  | Test 2 |  | N |  |
|  |  | Test 3 |  | *A. baumannii* |  |
| C24 |  | Test 1 |  | N |  |
|  |  | Test 2 |  | N |  |
| C26 |  | Test 1 |  | *A. baumannii* |  |
|  |  | Test 2 |  | *A. baumannii* |  |
|  |  | Test 3 |  | *A. baumannii* |  |
| C28 |  | Test 1 |  | *S. aureus* |  |
|  |  | Test 2 |  | N |  |
|  |  | Test 3 |  | *A. baumannii* |  |
| C30 |  | Test 1 |  | N |  |
|  |  | Test 2 |  | *A. baumannii* |  |
|  |  | Test 3 |  | *A. baumannii* |  |
| C32 |  | Test 1 |  | N |  |
|  |  | Test 2 |  | *K. pneumoniae* |  |
|  |  | Test 3 |  | *A. baumannii*  *K. pneumoniae* |  |
| C34 |  | Test 1 |  | N |  |
| C36 |  | Test 1 |  | *S. aureus* |  |
|  |  | Test 2 |  | *A. baumannii* |  |
|  |  | Test 3 |  | *A. baumannii* |  |
| C38 |  | Test 1 |  | *A. baumannii* |  |
|  |  | Test 2 |  | *A. baumannii* |  |
|  |  | Test 3 |  | *A. baumannii* |  |
| C40 |  | Test 1 |  | *P. aeruginosa* |  |
|  |  | Test 2 |  | *P. aeruginosa* |  |
| C42 |  | Test 1 |  | N |  |
|  |  | Test 2 |  | N |  |
| C44 |  | Test 1 |  | *S. aureus* |  |
|  |  | Test 2 |  | *A. baumannii* |  |
| C46 |  | Test 1 |  | N |  |
|  |  | Test 2 |  | N |  |
|  |  | Test 3 |  | *A. baumannii* |  |
| C48 |  | Test 1 |  | *A. baumannii* |  |
|  |  | Test 2 |  | *A. baumannii* |  |

**Supplementary Table 5: Demographic and baseline characteristics of the patients in the validation stage II.**

|  | **Experimental group (n=22)** | **Control group**  **(n=24)** | **p value** |
| --- | --- | --- | --- |
| Women | 9 (40.9%) | 11 (45.8%) | 0.774 |
| Men | 13 (59.1%) | 13 (54.2%) | 0.774 |
| Age, years (SD) | 58 (17.4) | 68 (9.5) | 0.015 |
| **Patients****' numbers of chronic comorbidities** | | |  |
| Hypertension | 9 (40.9%) | 17 (70.8%) | 0.073 |
| Coronary artery disease | 1 (4.5%) | 3 (12.5%) | 0.609 |
| Chronic pulmonary disease | 2 (9.1%) | 4 (16.7%) | 0.667 |
| Chronic kidney disease | 2 (9.1%) | 6 (25.0%) | 0.247 |
| Diabetes | 5 (22.7%) | 12 (50.0%) | 0.072 |
| Malignancy | 0 (0.0%) | 2 (8.3%) | 0.490 |
| Stroke | 3 (13.6%) | 8 (33.3%) | 0.171 |
| Immunodeficiency/immune suppressive therapy | 5 (22.7%) | 3 (12.5%) | 0.451 |
| Recent surgery | 4 (18.2%) | 3 (12.5%) | 0.694 |
| **Hemodynamic support (using vasoactive drugs)** | 7 (31.8%) | 7 (29.2%) | 1.000 |
| Norepinephrine ≤ 0.1 μg/(kg•min) | 2 | 3 |  |
| Norepinephrine > 0.1 μg/(kg•min) | 1 | 1 |  |
| Dopamine ≤ 5 μg/(kg•min) | 3 | 2 |  |
| Dopamine > 5 μg/(kg•min) | 0 | 1 |  |
| Dobutamine ≤ 5 μg/(kg•min) | 1 | 0 |  |
| Dobutamine > 5 μg/(kg•min) | 0 | 0 |  |
| **Status at randomization (D1)** | | |  |
| Temperature, °C | 38.4 (0.6) | 38.3 (0.7) | 0.345 |
| Coma | 6 (27.3%) | 6 (25.0%) | 1.000 |
| Systolic blood pressure, mmHg | 112.2 (19.3) | 121.6 (17.8) | 0.057 |
| Invasive mechanical ventilation | 20 (90.9%) | 24 (100.0%) | 0.223 |
| Renal replacement therapy | 0 (0.0%) | 2 (8.3%) | 0.493 |
| SOFA score | 6.3 (0.7) | 6.0 (0.5) | 0.935 |
| APACHE II score | 17.3 (1.6) | 18.9 (1.5) | 0.422 |
| Albumin, g/L | 32.1 (5.1) | 31.0 (3.7) | 0.442 |
| Globulin, g/L | 21.8 (3.9) | 23.1 (5.9) | 0.489 |
| Absolute lymphocyte count, 10^9^/L | 0.9 (0.7) | 0.7 (0.3) | 0.909 |
| White blood cells, 10^9^/L | 11.0 (5.7) | 12.7 (6.3) | 0.210 |
| CRP, mg/L | 94.7 (101.2) | 108.3 (84.0) | 0.424 |

SOFA score and APACHE II score are mean (SEM), other data are mean (SD), n (%). Mean (SEM/SD) is compared using Mann-Whitney test, and n (%) is compared using Fisher's exact test.

**Supplementary Table 6: Patients****' clinical outcomes.**

|  | **Experimental group (n=22)** | **Control group**  **(n=24)** | **p** |
| --- | --- | --- | --- |
| **Number of patients who have clinical indexes improved** | | | |
| Day 3 vs. Day 1 | | | |
| Temperature, °C | 15 (68.2%) | 9 (37.5%) | 0.045 |
| WBC, 10^9^/L | 15 (68.2%) | 12 (50.0%) | 0.211 |
| PCT, ng/mL | 18 (81.8%) | 19 (82.6%) | 0.945 |
| Day 7 vs. Day 1 | | | |
| Temperature, °C | 13 (72.2%) | 12 (54.5%) | 0.332 |
| WBC, 10^9^/L | 16 (84.2%) | 11 (50.0%) | 0.021 |
| PCT, ng/mL | 13 (68.4%) | 19 (86.4%) | 0.166 |
| Day 10 vs. Day 1 | | | |
| Temperature, °C | 13 (82.6%) | 12 (70.6%) | 0.688 |
| WBC, 10^9^/L | 9 (56.3%) | 8 (47.1%) | 0.598 |
| PCT, ng/mL | 12 (75.0%) | 15 (88.2%) | 0.325 |
| **Number of patients undergoing effective treatment** | | | |
| Day 3 | 13 (59.1%) | 11 (45.8%) | 0.395 |
| Day 7 | 16 (84.2%) | 11 (50.0%) | 0.046 |
| Day 10 | 13 (81.3%) | 10 (58.8%) | 0.259 |
| **Clinical endpoint outcomes** | | | |
| 28-days mortality | 8 (36.4%) | 8 (33.3%) | 1.000 |
| Mechanical ventilation from randomization to 28th day, days | 11.3 (7.7) | 11.5 (7.7) | 0.970 |
| Shock from randomization to 28th day, days | 3.1 (4.3) | 2.3 (3.6) | 0.456 |
| Numbers of antibiotic-associated diarrhea | 0 (0.0%) | 2 (8.3%) | 0.490 |

For those data are n (%), all p values are calculated using Fisher's exact tests. For those data are mean (SD), all p values are calculated using Mann-Whitney tests.

**Supplementary Table 7: Potential competitive analysis among bacteria.**

| **Sample ID** | **Bacteria detected by SSBD grow**  **in CCT tests** | **Bacteria detected by SSBD could not grow**  **in CCT tests** | **Probable relations**  **among bacteria** |
| --- | --- | --- | --- |
| A05 | *K. pneumoniae* (III)  *P. aeruginosa* (III) | *A. baumannii* (III) | *K. pneumoniae* + *P. aeruginosa*  > *A. baumannii* |
| A09 | *A. baumannii* (II) | *S. aureus* (I) | Strength: II > I |
| A10 | *A. baumannii* (III)  *K. pneumoniae* (III) | *P. aeruginosa* (I) | Strength: III + III > I |
| A11 | *A. baumannii* (III) | *K. pneumoniae* (I) | Strength: III > I |
| A16 | *K. pneumoniae* (III) | *P. aeruginosa* (III) | *K. pneumoniae* > *P. aeruginosa* |
| A21 | *A. baumannii* (III)  *S. aureus* (I) | *K. pneumoniae* (III) | Strength: III + I > III |
| A22 | *A. baumannii* (III) | *S. epidermidis* (III) | *A. baumannii* > *S. epidermidis* |
| A27 | *A. baumannii* (III) | *S. epidermidis* (III) | *A. baumannii* > *S. epidermidis* |
| A28 | *A. baumannii* (II) | *P. aeruginosa* (III) | *A. baumannii* > *P. aeruginosa* |
| A30 | *A. baumannii* (III) | *S. aureus* (III)  *S. epidermidis* (I) | *A. baumannii*  > *S. aureus* + *S. epidermidis* |
| A33 | *S. aureus* (I) | *S. maltophilia* (II) | *S. aureus* > *S. maltophilia* |
| A36 | *A. baumannii* (III) | *S. maltophilia* (I) | *A. baumannii* > *S. maltophilia* |
| A37 | *A. baumannii* (III) | *S. epidermidis* (III) | *A. baumannii* > *S. epidermidis* |
| A45 | *A. baumannii* (III) | *K. pneumoniae* (II) | *A. baumannii* > *K. pneumoniae* |
| A49 | *A. baumannii* (III)  *S. aureus* (III) | *S. maltophilia* (III) | *A. baumannii* + *S. aureus*  > *S. maltophilia* |
| A55 | *A. baumannii* (III) | *E. coli* (II)  *K. pneumoniae* (III)  *S. maltophilia* (III) | *A. baumannii* > *E. coli* +*K. pneumoniae* + *S. maltophilia* |
| A65 | *A. baumannii* (III)  *P. aeruginosa* (I) | *S. maltophilia* (II) | Strength: III + I > II |
| A68 | *A. baumannii* (III) | *P. aeruginosa* (III) | *A. baumannii* > *P. aeruginosa* |
| A69 | *A. baumannii* (I) | *P. aeruginosa* (III) | *A. baumannii* > *P. aeruginosa* |
| A72 | *A. baumannii* (III) | *S. maltophilia* (I) | *A. baumannii* > *S. maltophilia* |
| A73 | *A. baumannii* (III)  *P. aeruginosa* (III) | *K. pneumoniae* (III) | *A. baumannii* + *P. aeruginosa*  > *K. pneumoniae* |
| A75 | *A. baumannii* (III) | *P. aeruginosa* (III) | *A. baumannii* > *P. aeruginosa* |
| A76 | *P. aeruginosa* (III) | *A. baumannii* (I)  *S. maltophilia* (I) | Strength: III > I + I |
| B05-1 | *A. baumannii* (III) | *S. maltophilia* (II) | *A. baumannii* > *S. maltophilia* |
| B05-2 | *A. baumannii* (III) | *S. maltophilia* (I) | *A. baumannii* > *S. maltophilia* |
| B09-1 | *A. baumannii* (III) | *S. aureus* (III)  *S. capitis* (III)  *S. maltophilia* (III) | *A. baumannii* > *S. aureus*  + *S. capitis* + *S. maltophilia* |
| B11-1 | *A. baumannii* (III) | *P. aeruginosa* (III)  *S. maltophilia* (II) | *A. baumannii*  > *P. aeruginosa* + *S. maltophilia* |
| B17-3 | *A. baumannii* (I) | *K. pneumoniae* (I)  *E. faecalis* (I) | *A. baumannii*  > *K. pneumoniae* + *E. faecalis* |
| B19-1 | *A. baumannii* (III) | *P. aeruginosa* (I)  *S. maltophilia* (II) | Strength: III > I + II |
| B19-2 | *A. baumannii* (III), *S. maltophilia* (III) | *P. aeruginosa* (III) | *A. baumannii* + *S. maltophilia*  > *P. aeruginosa* |
| B19-3 | *K. pneumoniae* (III), *S. maltophilia* (III) | *P. aeruginosa* (III) | *K. pneumoniae* + *S. maltophilia*  > *P. aeruginosa* |
| B21-1 | *S. aureus* (III)  *S. maltophilia* (III) | *P. aeruginosa* (III) | *S. maltophilia* + *S. aureus*  > *P. aeruginosa* |
| B21-2 | *S. maltophilia* (III) | *S. aureu* (III), *P. aeruginosa* (III) | *S. maltophilia* > *S. aureus* + *P. aeruginosa* |
| B23-1 | *S. maltophilia* (III) | *S. capitis* (I) | Strength: III > I |
| B31-1 | *A. baumannii* (III) | *S. capitis* (III) | *A. baumannii* > *S. capitis* |
| B39-2 | *K. pneumoniae* (III)  *P. aeruginosa* (III) | *A. baumannii* (I)  *E. coli* (I) | Strength: III + III > I + I |
